# Influence of sexual risk behaviour and STI co-infection dynamics on the evolution of HIV set point viral load in MSM

**DOI:** 10.1101/19011221

**Authors:** Diana M Hendrickx, Wim Delva, Niel Hens

**Affiliations:** I-BioStat, Data Science Institute, Hasselt University, Hasselt, Belgium; The South African Department of Science and Technology-National Research Foundation (DST-NRF) Centre of Excellence in Epidemiological Modelling and Analysis (SACEMA), Stellenbosch University, Stellenbosch, South Africa; Department of Global Health, Faculty of Medicine and Health, Stellenbosch University, Stellenbosch, South Africa; International Centre for Reproductive Health, Ghent University, Ghent, Belgium; Rega Institute for Medical Research, KU Leuven, Leuven, Belgium; School for Data Science and Computational Thinking, Stellenbosch University, Stellenbosch, South Africa; Centre for Health Economics Research and Modelling Infectious Diseases, Vaccine & Infectious Disease Institute, University of Antwerp, Antwerp, Belgium

## Abstract

HIV viral load (VL) is an important predictor of HIV progression and transmission. Anti-retroviral therapy (ART) has been reported to reduce HIV transmission by lowering VL. However, apart from this beneficial effect, increased levels of population mean set-point viral load (SPVL), an estimator for HIV virulence, have been observed in men who have sex with men (MSM) in the decade following the introduction of ART in the Netherlands. Several studies have been devoted to explain these counter-intuitive trends in SPVL. However, to our knowledge, none of these studies has investigated an explanation in which it arises as the result of a sexually transmitted infection (STI) co-factor in detail.

In this study, we adapted an event-based, individual-based model to investigate how STI co-infection and sexual risk behaviour affect the evolution of HIV SPVL in MSM before and after the introduction of ART.

The results suggest that sexual risk behaviour has an effect on SPVL and indicate that more data are needed to test the effect of STI co-factors on SPVL. Furthermore, the observed trends in SPVL cannot be explained by sexual risk behaviour and STI co-factors only.

We recommend to develop mathematical models including also factors related to viral evolution as reported earlier in the literature. However, this requires more complex models, and the collection of more data for parameter estimation than what is currently available.

## 1 Introduction

HIV viral load (HIV RNA concentration in plasma) is an important and widely used prognostic marker for HIV disease progression and transmission [24, 27, 36]. While immediately after infection rapid HIV replication and high HIV viral load (VL) values are observed, VL declines during the asymptomatic phase and reaches a stable level, called set-point viral load (SPVL) after a few weeks to a few months [13]. Evolution of HIV virulence is often estimated by means of the evolution of the population mean SPVL [22].

It has been reported that anti-retroviral therapy (ART) reduces HIV transmission by decreasing VL [28, 31], and that no HIV transmission happens under successful ART. However, apart from these beneficial effects, increased population mean SPVL levels, corresponding to increased HIV virulence, were observed in MSM in the Netherlands during the period 1995-2007, after the introduction of ART in 1994 [19]. Moreover, an increase in the number of new HIV cases in MSM in the Netherlands has been observed during the period 1996-2008. After 2008, a drop in new HIV diagnoses has been observed [45].

Several studies have been devoted to explain these counter-intuitive trends in HIV SPVL after the introduction of ART. The majority of these studies try to explain these trends from an evolutionary perspective. They explain the observed trends by host genetic factors [43, 52], viral adaptation of HIV to its host population [5, 8, 14, 38] and evolution of HIV SPVL to maintain higher viral fitness [12, 18, 39, 42]. HIV SPVL has been reported to be partially heritable from the infected donor to the susceptible recipient during HIV transmission [2, 12]. This has led to the hypothesis that sexual risk behaviour could influence the evolution of SPVL [11, 17]. A few studies have investigated sexual risk behaviour as a potential factor influencing SPVL [1, 17]. A recent study of Goodreau et al. [17] reports a positive relationship between relational concurrency and mean SPVL.

Mathematical modeling has been shown to be a valuable tool to investigate the evolution of SPVL and HIV virulence. Bezemer et al. [1] used a deterministic compartmental ordinary differential equation (ODE) model to study the influence of sexual risk behaviour on the HIV epidemic. Herbeck et al. [23] used an individual-based model to investigate trends in HIV virulence and community viral load. Their model contained functions for transmission and progression of HIV, SPVL, VL at multiple stages of HIV and SPVL heritability. Sexual relationships were described by a simplified contact network, without including sexual mixing patterns. Roberts et al. [33] designed a deterministic model to assess the impact of ART on viral evolution. Smith et al. [40] developed a compartmental model to study the influence of ART and pre-exposure prophylaxis (PrEP) on the evolution of HIV virulence. Goodreau et al. [17] used an individual- and network-based model to study the relationship between relational concurrency, HIV stages and evolution of HIV SPVL. However, none of these models implemented the effect of a sexually transmitted infection (STI) co-factor whereas the presence of an STI co-factor has been suggested to influence the trend in SPVL [19].

HIV-negative people infected with an STI are reported to have a higher risk for HIV acquisition, while HIV-positive people co-infected with an STI have a higher risk of transmitting HIV to another individual [47]. As a consequence, a higher incidence of an STI would lead to an increased number of new HIV patients. Furthermore, an observational study of Scheer et al [37] in MSM reported that people on ART show an increased risk of developing an STI, and this effect occurs most likely as a consequence of sexual risk behaviour. Moreover, having HIV also increases the risk for acquisition of an STI [26]. Furthermore, Gras et al. [19] observed a positive correlation between SPVL levels and the number of HIV patients. This has led to the hypothesis that the presence of an STI co-factor could influence the evolution of SPVL by increasing the number of new HIV cases.

In this study, we adapted an event-based, individual-based model to investigate the effect of sexual risk behaviour, STI co-infection and their combined effect on the evolution of HIV SPVL in MSM after the introduction of ART.

## 2 Materials and Methods

### 2.1 Simpact Cyan 1.0 modeling framework

Simpact Cyan 1.0 is a open-source framework for constructing individual-based models for simulating the transmission, diagnosis and treatment of HIV [29]. The program models each individual in a heterosexual population or a population of MSM, and the sexual relationships between individuals. The formation and dissolution of relationships, as well as birth, mortality, HIV transmission, diagnosis and treatment are represented by events, which have a certain risk of taking place at a certain moment, represented by their hazard function. Models are implemented in continuous time and updated each time an event happens. Simpact Cyan 1.0 also implements a generic sexually transmitted infection (STI) co-factor effect on HIV. Furthermore, it is also possible to simulate interventions by changing certain parameters during the simulation.

More detailed information on the Simpact Cyan 1.0 modeling framework is available in [29] and from http://www.simpact.org/.

### 2.2 Implementation of viral load and set-point viral load

When an individual becomes infected by HIV, a value for the SPVL is assigned to this individual. SPVL values on a base 10 logaritmic scale were drawn from a binomial distribution using the same mean and standard deviation in both dimensions:

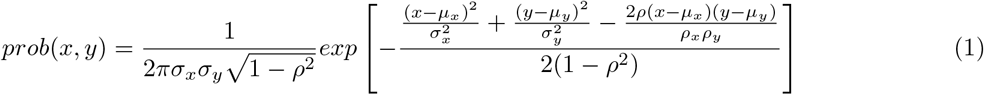

The two dimensions represent the persons that are already infected (x) and the persons becoming infected (y). The newly infected individual’s SPVL depends on the SPVL of the infector.

*µ*_*x*_, *µ*_*y*_: mean of the distribution of SPVL

*σ*_*x*_, *σ*_*y*_: standard deviation of the distribution of SPVL

*ρ*: correlation between x and y in the calculation of SPVL

Parameter *ρ* represents SPVL heritability and was taken from Blanquart et al. [2]. The mean and standard deviation were fitted to the histogram for MSM in Fraser et al. [11]. The values of the parameters are provided in Table S1.

The SPVL is the VL of a person during the chronic stage. In the acute stage, AIDS stage and final AIDS stage, the VL differs from the SPVL such that the HIV transmission hazard increases by a factor 10, 7 and 12 respectively. These multipliers are the default parameters in Simpact Cyan 1.0. They are based on a previous model fit [29] to data generated by Fraser et al. [11].

### 2.3 Implementation of treatment with anti-retroviral therapy

After being infected with HIV, an individual gets diagnosed as being HIV positive through a diagnosis event. In this study the hazard for diagnosis depends on a single baseline parameter: *hazard* = *exp*(*baseline*).

After being diagnosed, the person’s progression of HIV is monitored by inspecting his CD4 count during an HIV monitoring event. A person is eligible to be treated with ART if his CD4 count is below a given threshold.

In this study, we assume ART is offered to all eligible individuals and that all persons will accept treatment when offered. This is implemented by setting the ART acceptance threshold equal to 1 (see Table S1). Furthermore, we assume that no ART treatment is available before 1994, in correspondence with Palella et al. [32]. To implement that nobody will be treated before 1994, we set the hazard for diagnosis equal to 0 (*exp*(*baseline*) = 0). From 1994 to the end of the simulation, availability of ART in increased by

- increasing the CD4 threshold from 200 in 1994 to 500 in 2014 with steps of 30 in every two years (see Table S2);
- increasing the baseline parameter for the diagnosis hazard from −1.5 in 1994 to 1 in 2014 with steps of 0.25 in every two years (see Table S2).

When an individual starts with ART, his current VL decreases by 70% (default in Simpact Cyan 1.0).

### 2.4 Implementation of the STI co-factor effect

The start of the STI, in this study HSV-2, is implemented as an HSV-2 seeding event, where a certain amount of MSM are marked as HSV-2 infected. Transmission of the STI is described by the HSV-2 transmission hazard:

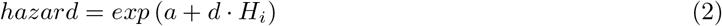

In equation (2), the parameter a provides a baseline value for HSV-2 transmission. The parameter d describes the effect of HIV on HSV-2 transmission. *H*_*i*_ is a binary indicator for the HIV status of the HSV-2 infected person: 1 = HIV-positive, 0 = HIV-negative.

The parameters for HSV-2 seeding and transmission were taken from the literature and are provided in Table S1 of the Supplementary Material.

The effects of the STI on HIV are modeled within the HIV transmission hazard:

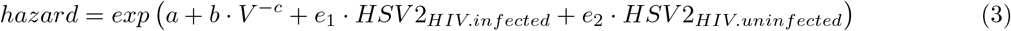

In equation (3), the parameter a provides a baseline value for HIV transmission. Parameters b and c describe the influence of the viral load (V) of the HIV infected person on HIV transmission. Parameters *e*_1_ and *e*_2_ describe the effect of the STI on HIV transmission. Parameter *e*_1_ represents the higher risk of an HIV-positive person co-infected with HSV-2 to transmit HIV to another person. Parameter *e*_2_ represents the higher risk of a HIV-negative person infected with HSV-2 to acquire HIV. *HSV* 2_*HIV*.*infected*_ and *HSV* 2_*HIV*.*uninfected*_ are binary indicators for the HSV-2 status of the HIV-infected and HIV-uninfected person respectively (1 = HSV-2-positive, 0 = HSV-2-negative).

The values of the HIV transmission parameters a, b and c are determined during the calibration procedure described in paragraph 2.8. The formula for a hazard depending on viral load is based on a study by Hargrove et al. [21]. Parameters *e*_1_ and *e*_2_ were taken from the literature and are provided in Table S1 of the Supplementary Material.

### 2.5 Implementation of sexual behaviour

In Simpact Cyan 1.0, sexual behaviour among MSM is described by MSM formation and dissolution events, which are described by their hazard function. According to previous studies, relationship formation is influenced by

- the number of relationships each partner has [17] and its assortativity [35];
- the age of the partners [30, 34] and its assortativity [49].

In this study, the hazard for relationship formation is described by the following formula, where the selection of the parameters is based on the previous studies described above:

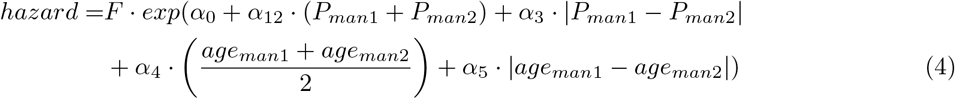

In equation (4), F is a normalization factor, which roughly divides the formation hazard by the population size. Applying this normalization factor avoids that an increase in population size automatically results in more relationships. *P*_*man*1_ and *P*_*man*2_ represent the number of partners the two men already have; *age*_*man*1_ and *age*_*man*2_ represent the age of the two men. Parameter *α*_0_ represents a baseline value for the formation hazard. *α*_12_ describes the influence of the number of relationships the men already have, while *α*_3_ captures its assortativity by adding a weight for the effect of the difference in number of partners, between the two men on relationship formation. *α*_12_ ranges from minus infinity (-inf) to 0, where -inf corresponds to a population where everyone is monogamous, while 0 corresponds with no influence of the number of partners. The more negative, the less concurrent partnerships are formed. E.g. if this parameter is equal to −5, this means that the hazard is multiplied with exp(−5) (or in other words divided by exp(5)) if one of the men already has a partner. The parameter *α*_3_ is 0 in case of no assortativity. A more negative parameter corresponds to more assortativity. E.g. if this parameter is equal to log(0.5), then the formation hazard decreases with 50% if the number of current partners between the two men differs 1. *α*_4_ describes the influence of the mean age of the two men, while *α*_5_ captures its assortativity by adding a weight for the importance of the difference in age between the two men when forming a relationship. A negative value of *α*_4_ means that older persons form less relationships, while a positive value means that more relationships are formed by older persons. Both have been reported in the literature, e.g. in a study of Marcus et al. [30] older persons have less partners, while in another study conducted by Rozenberg et al. [34] less partners were reported for younger people. Therefore, both positive and negative values for parameter *α*_4_ were explored in this study. In case of no assortativity, *α*_5_ is 0. A more negative parameter corresponds with more assortativity. E.g. *α*_5_ = *log*(0.5) corresponds with a decrease of the formation hazard with 50% if the age between the two men differs 1 year.

The values of the parameters *α*_0_, *α*_12_, *α*_3_, *α*_4_, *α*_5_ are determined during the calibration procedure described in paragraph 2.8.

In this study, the break-up of relationships is described by the dissolution hazard:

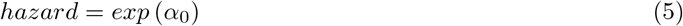

The value of the parameter *α*_0_ in equation (4) is determined during the calibration procedure described in paragraph 2.8.

### 2.6 Implementation of a change in risk behaviour with increasing availability of ART

In absence of a change in risk behaviour, the parameter *α*_12_ in de formation hazard (equation (4)) is constant over the whole simulation period.

A change in risk behaviour with increasing availability of ART is implemented by increasing the value of *α*_12_ with 0.05 every two years between 1994 and 2014, like represented in Table S2 of the Supplementary Material. The initial value of *α*_12_ is determined during the calibration procedure described in paragraph 2.8.

### 2.7 Model scenarios

In this study, we focus on the influence of sexual risk behaviour and STI co-infection dynamics on the evolution of the HIV set point viral load (SPVL) in MSM. Simulations are conducted for the period from 1980 to 2015, including both the period before and after the introduction of ART in 1994. After the introduction of ART, it is also assumed that the CD4 count threshold below which ART is offered increased [9], so that more people are treated. Furthermore, we assume that availability of ART has led to intensified HIV testing [15].

Four model scenarios including ART coverage are considered:

- no STI co-factor effect and no change in risk behaviour (nSTI-nBC): null scenario;
- STI co-factor effect (implemented as described in paragraph 2.4.) and no change in risk behaviour (STI-nBC);
- a change in risk behaviour implemented as described in paragraph 2.5.) and no STI co-factor effect (nSTI-BC);
- a change in risk behaviour (implemented as described in paragraph 2.5.) and an STI co-factor effect (implemented as described in paragraph 2.4.)(STI-BC).

In case any of the other scenarios has an improved GOF to the SPVL data compared to the null model, and/or better describes the SPVL trends, this suggests that the factors included in this scenario (STI co-factor and/or behavioural change) contribute to explaining the SPVL trends.

Parameters based on prior knowledge used in the model simulations, and intervention events for simulating the increase in ART coverage since 1994, and the increased risk behaviour with ART coverage (if applicable) are described in the Supplementary Material.

### 2.8 Model calibration

Table 1 presents the available literature data on the HIV epidemic in MSM in the Netherlands, used for model calibration. SPVL and HIV prevalence data are from different, but overlapping time intervals (SPVL: 1985-2007, HIV prevalence: 2003-2012).

**Table 1.**
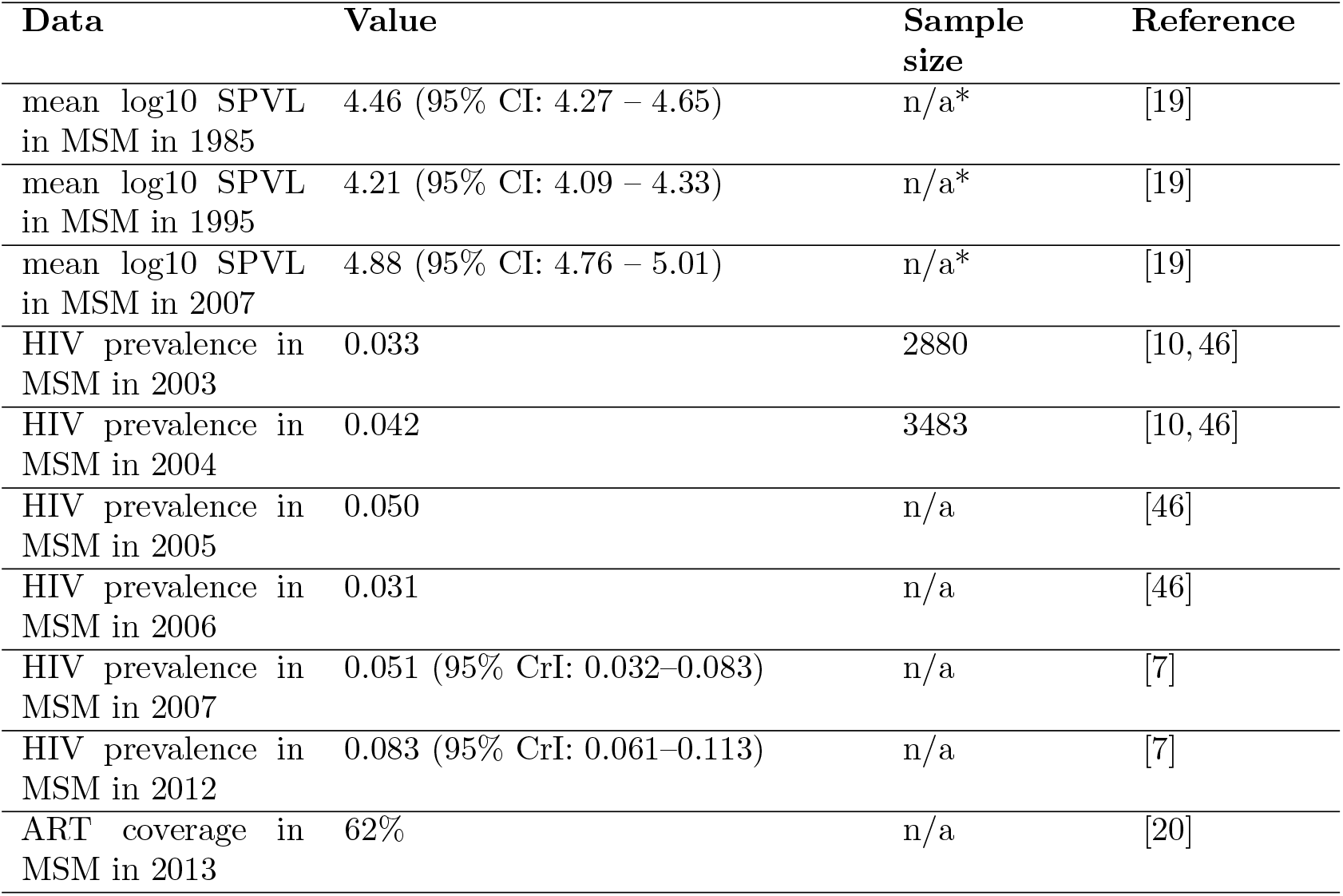
Available literature data used for model calibration. All data are for the Netherlands. CI: confidence interval; CrI: credible interval. * The values of the mean log10 SPVL in Gras et al. [19] were obtained from data of the mean HIV-1 RNA concentration at set-point of 612 MSM with date of seroconversion between 1985 and 2007. Gras et al. fitted a cubic spline through these data and estimated values for 1985, 1995 and 2007 from this spline.

| Data | Value | Sample size | Reference |
| --- | --- | --- | --- |
| mean log10 SPVL in MSM in 1985 | 4.46 (95% CI: 4.27 – 4.65) | n/a* | [19] |
| mean log10 SPVL in MSM in 1995 | 4.21 (95% CI: 4.09 – 4.33) | n/a* | [19] |
| mean log10 SPVL in MSM in 2007 | 4.88 (95% CI: 4.76 – 5.01) | n/a* | [19] |
| HIV prevalence in MSM in 2003 | 0.033 | 2880 | [10, 46] |
| HIV prevalence in MSM in 2004 | 0.042 | 3483 | [10, 46] |
| HIV prevalence in MSM in 2005 | 0.050 | n/a | [46] |
| HIV prevalence in MSM in 2006 | 0.031 | n/a | [46] |
| HIV prevalence in MSM in 2007 | 0.051 (95% CrI: 0.032–0.083) | n/a | [7] |
| HIV prevalence in MSM in 2012 | 0.083 (95% CrI: 0.061–0.113) | n/a | [7] |
| ART coverage in MSM in 2013 | 62% | n/a | [20] |

For the nine parameters in Table 2, there were no literature values available. Therefore, these parameters were fitted to the data in Table 1 by applying an active learning approach [50] using the following steps:

**Table 2.**
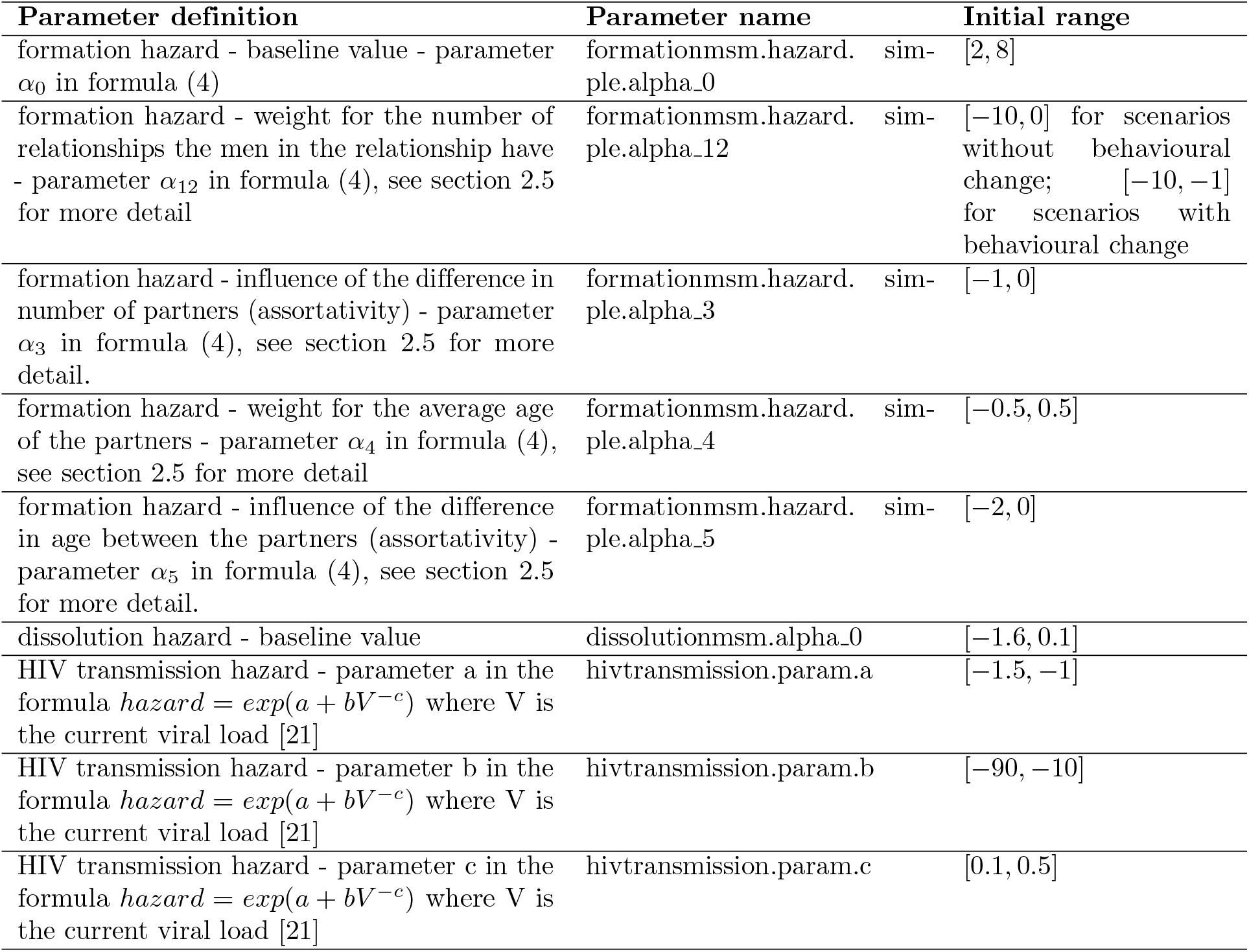
Parameters fitted to the data in Table 1. Initial ranges are based on the examples in the RSimpactHelper package https://github.com/wdelva/RSimpactHelp. The parameter names in the second column are directly related to Simpact Cyan 1.0.

1. Select 10,000 parameter sets by applying Latin Hypercube Sampling (LHS) [41] and using the initial parameter ranges from Table 2.
2. For each of the 10,000 parameter sets, run a simulation with Simpact Cyan 1.0.
3. For each simulation, calculate the goodness-of-fit (GOF) based on the sum of squared relative errors [4].
4. Apply the selection procedure of Castro Sanchez et al [3] based on the GOF measure to narrow the solution space (intervals for the parameters).
5. Repeat steps 1-4 using the new intervals for the parameters until the GOF does not improve anymore.

When selecting the final parameters, it was also taken into account that the results show realistic point prevalence of partnership concurrency. Unfortunately, no estimates of partnership concurrency were reported for MSM in the Netherlands. Studies in the US report estimates of partnership concurrency equal to 14.8% [16] and 26.3% [48] for MSM. Based on these two studies, we assume that realistic values for the point prevalence of concurrency are between 10% and 30%.

Apart from the overall GOF, also the GOF based on SPVL data only, the GOF based on HIV prevalence data only, and the GOF to the ART coverage of 2013 are reported.

The HIV transmission hazard: *hazard* = *exp*(*a* + *bV* ^−*c*^+ additional terms), where V is the current viral load, is based on equation (9) of Hargrove et al. [21]. Its default parameters (*a* = *—*1.3997, *b* = − 12.0220, *c* = 0.1649) are obtained by fitting parameters a, b and c to the data from Fraser et al. [11]. In this study, additional parameters are added to this hazard in case an STI co-factor is simulated (see paragraph 2.4).

## 3 Results

### 3.1 Model calibration

Table 3 shows the fitted parameters for the four model scenarios.

**Table 3.** Fitted parameters for the four model scenarios. nSTI-nBC: no STI co-factor, no behavioural change; STI-nBC: STI co-factor, no behavioural change; nSTI-BC: no STI co-factor, behavioural change; STI-BC: STI co-factor, behavioural change.

| parameter | nSTI-nBC | STI-nBC | nSTI-BC | STI-BC |
| --- | --- | --- | --- | --- |
| formation baseline $\alpha_0$ (formula (4)) | 7.16 | 3.52 | 3.45 | 3.60 |
| formation weight number of partners $\alpha_{12}$ (formula (4)) | -5.48 | -8.88 | -9.76 | -4.66 |
| formation weight assortativity number of partners $\alpha_3$ (formula (4)) | -0.623 | -0.216 | -0.842 | -0.617 |
| formation weight age $\alpha_4$ (formula (4)) | 0.0312 | 0.242 | 0.244 | 0.0738 |
| formation weight age assortativity $\alpha_5$ (formula (4)) | -0.0368 | -1.04 | -0.170 | -0.176 |
| dissolution baseline $\alpha_0$ (formula (5)) | -0.200 | -1.52 | -0.794 | -0.0363 |
| HIV transmission a (formula (3)) | -1.03 | -1.21 | -1.01 | -1.45 |
| HIV transmission b (formula (3)) | -13.1 | -20.8 | -37.7 | -13.1 |
| HIV transmission c (formula (3)) | 0.370 | 0.449 | 0.495 | 0.397 |

A detailed statistical analysis of the parameter space exploring

- which parameters are most influenced by the data;
- which regions contain a large amount of solutions with low relative sum of squared errors;
- which parameters show associations

is provided in the Supplementary Material.

### 3.2 Goodness-of-fit

Four goodness-of-fit (GOF) statistics are considered: overall sum of squared relative errors (Figure 1A), sum of squared relative errors to the SPVL data only (Figure 1B), sum of squared relative errors to the HIV prevalence data only (Figure 1C) and squared relative error to the ART coverage in 2013 (Figure 1D). Figure 1A shows that including an STI co-factor effect or/and a behavioural change results in a better overall goodness-of-fit (GOF) to the literature data. However, when considering only GOF to the three data points on SPVL, none of the models showed a better fit to the data than the model not including any of these two effects (see Figure 1B). The improved overall GOF for the models including an STI co-factor (STI-nBC) and/or a behavioural change (nSTI-BC) can be explained by the improved GOF to the HIV prevalence data (see Figure 1C). The model including both an STI co-factor and a behavioural change shows the best GOF to the HIV prevalence data, and also the best overall GOF.

**Figure 1.**
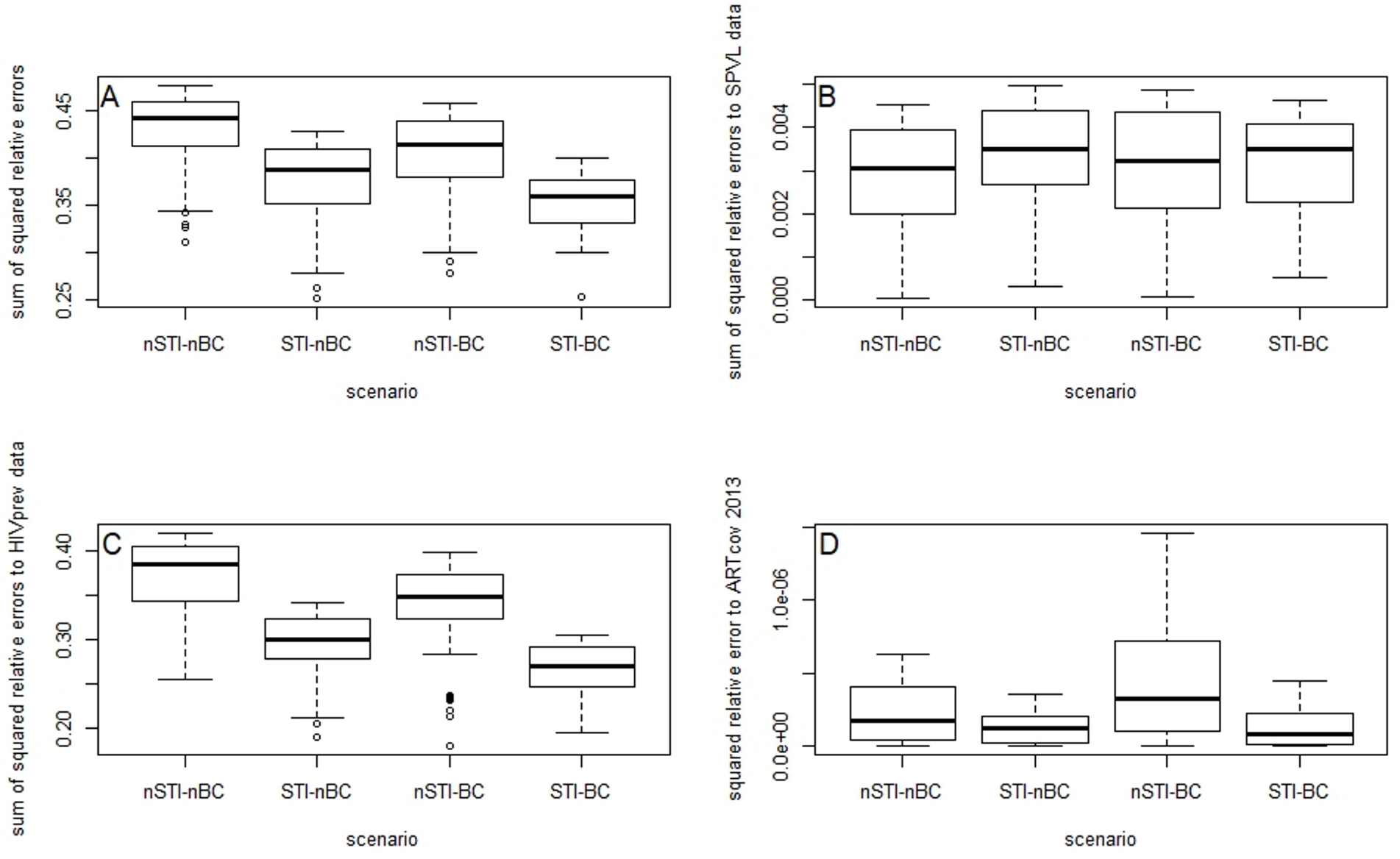
Boxplots for top 100 solutions based on overall goodness-of-fit. A) boxplots for overall sum of squared relative errors; B) boxplots for sum of squared relative errors to the SPVL data only; C) boxplots for sum of squared relative errors to the HIV prevalence data only; D) squared relative error to the ART coverage in 2013.

### 3.3 Trends in set-point viral load, HIV prevalence, ART coverage and HIV incidence

Figures 2 and 3A show the median population mean log10 SPVL and the interquartile range for 100 simulations with the four scenarios. The population mean log10 SPVL was calculated as the mean set-point HIV-RNA concentration at 9-27 months after seroconversion. None of the scenarios generates the increasing trend in population mean HIV SPVL between 1995 and 2007 observed by Gras et al [19]. In the nSTI-nBC (grey in Figure 3A) and nSTI-BC (blue in Figure 3A) scenarios (scenarios without STI co-factor), the mean log10 SPVL decreases from 1995 onwards. In the scenarios including an STI (STI-nBC (red in Figure 3A) and STI-BC (green in Figure 3A)), the mean log10 SPVL is constant between 1995 and 2000, and decreases from 2000 onwards.

**Figure 2.**
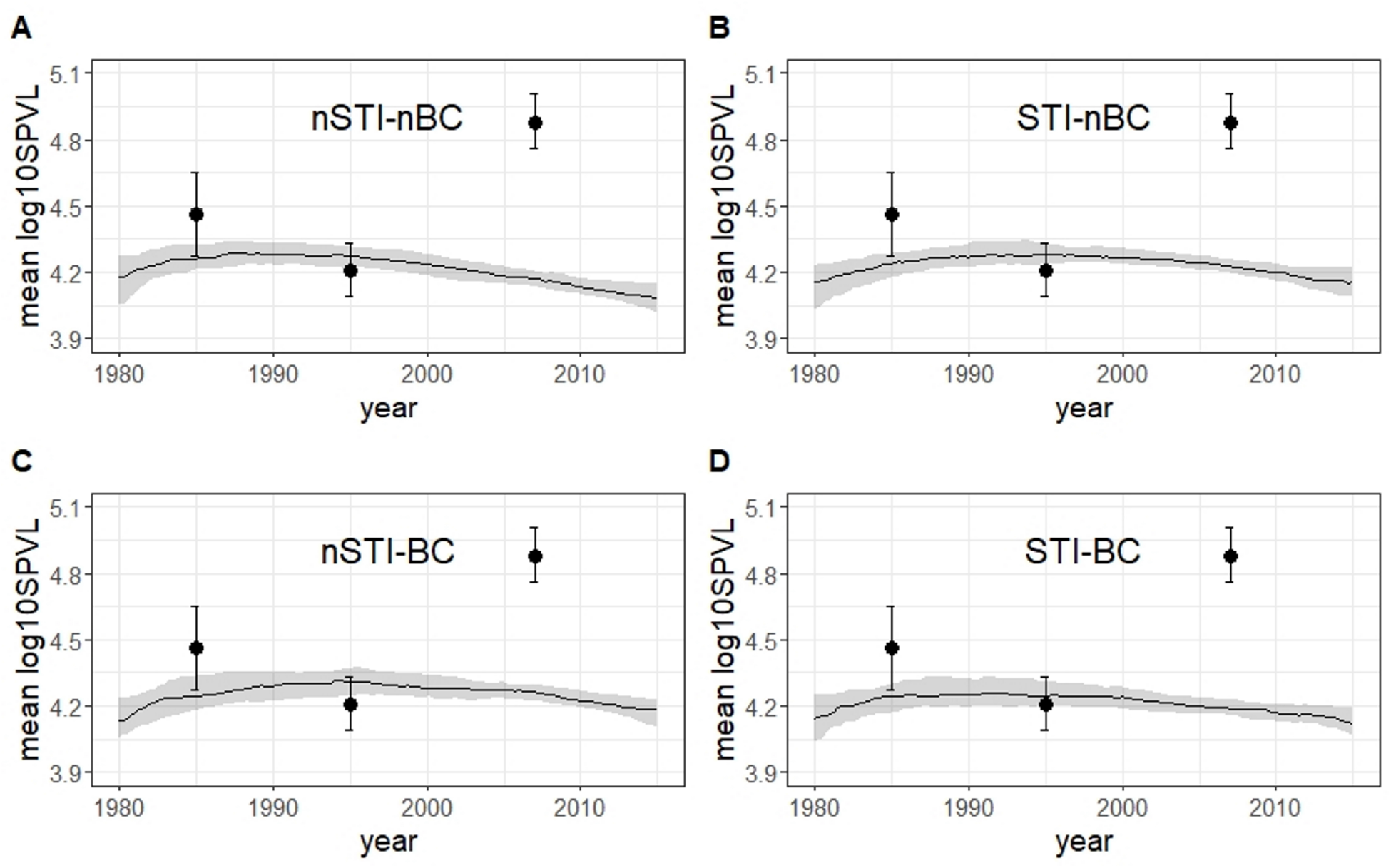
Mean log10 HIV SPVL (mean set-point HIV-RNA concentration at 9-27 months of untreated HIV infection). Median of 100 simulations with the fitted parameters (solid line) and interquartile range (shaded area) for the period 1980-2015. A: no STI co-factor, no behavioural change (nSTI-nBC); B: STI co-factor, no behavioural change (STI-nBC); C: no STI co-factor, behavioural change (nSTI-BC); D: STI co-factor, behavioural change (STI-BC). See Supplementary Material for more detail on how the figures were generated. The black dots and error bars represent the mean log10 SPVL with 95% confidence interval for 1985, 1995 and 2007 from [19].

**Figure 3.**
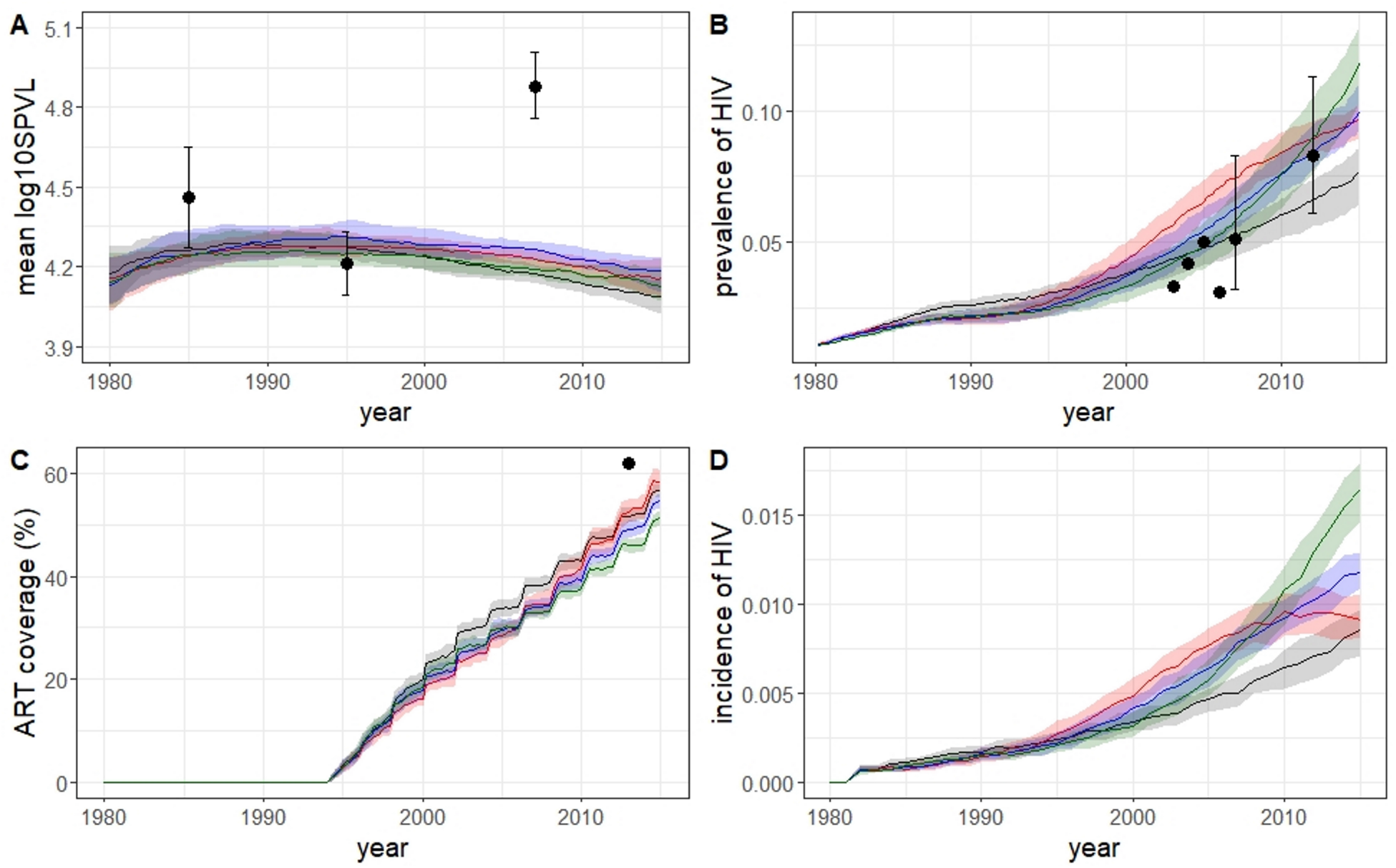
Mean log10 HIV SPVL (A), prevalence of HIV (B), ART coverage (C) and incidence of HIV (D) - median of 100 simulations with the fitted parameters (solid line) and interquartile range (shaded area) for the period 1980-2015. Grey: no STI co-factor, no behavioural change (nSTI-nBC); red: STI co-factor, no behavioural change (STI-nBC); blue: no STI co-factor, behavioural change (nSTI-BC); darkgreen: STI-cofactor, behavioural change (STI-BC). The black dots and the error bars represent the literature data from Table 1.

Figures 3B and C show how well the simulations of the four scenarios fit to the HIV prevalence and ART coverage data. All scenarios capture the increasing trend in HIV prevalence between 2003 and 2012. However, only the interquartile range for the STI-BC scenario includes the majority of the HIV prevalence data points from the period 2003-2007. In Figure 3C, we observe that the model scenario STI-nBC results in the closest estimate of the ART coverage of 62% in 2013 reported in [20]. Furthermore, for the STI-nBC scenario, also the year where the HIV incidence reaches its peak is the closest to the year of the peak observed in the literature data (2010 for STI-nBC, see Figure 3D; 2008 in the literature [45]). For all other scenarios, the HIV incidence increases until the end of the simulation (2015).

### 3.4 Relationship between HIV SPVL and point prevalence of concurrency

To investigate the relationship between HIV SPVL and points prevalence of concurrency, the weight for the number of partners (*α*_12_ in formula (4)) was varied between −10 and −1, while all other literature and estimated parameters were kept constant at their values in Table S1 and Table 3 respectively. A hundred simulations with values for *α*_12_ drawn from a uniform distribution with bounds −10 and −1 were performed. Figure 4 shows the mean log10 HIV SPVL of all individuals with a date of HIV seroconversion between 1980 and 2015 against mean point prevalence of concurrency over the period 1980-2015 for these 100 simulations with the four scenarios. All scenarios confirm the increasing trend of SPVL with relational concurrency described by Goodreau et al [17]. However, this increase in mean log 10 SPVL is small compared to the reported difference in mean log 10 SPVL between 1995 and 2007 (0.67), even when the point prevalence of concurrency changes from 0 to 1 (100%)(change of 0.10-0.15 of mean log 10 SPVL).

**Figure 4.**
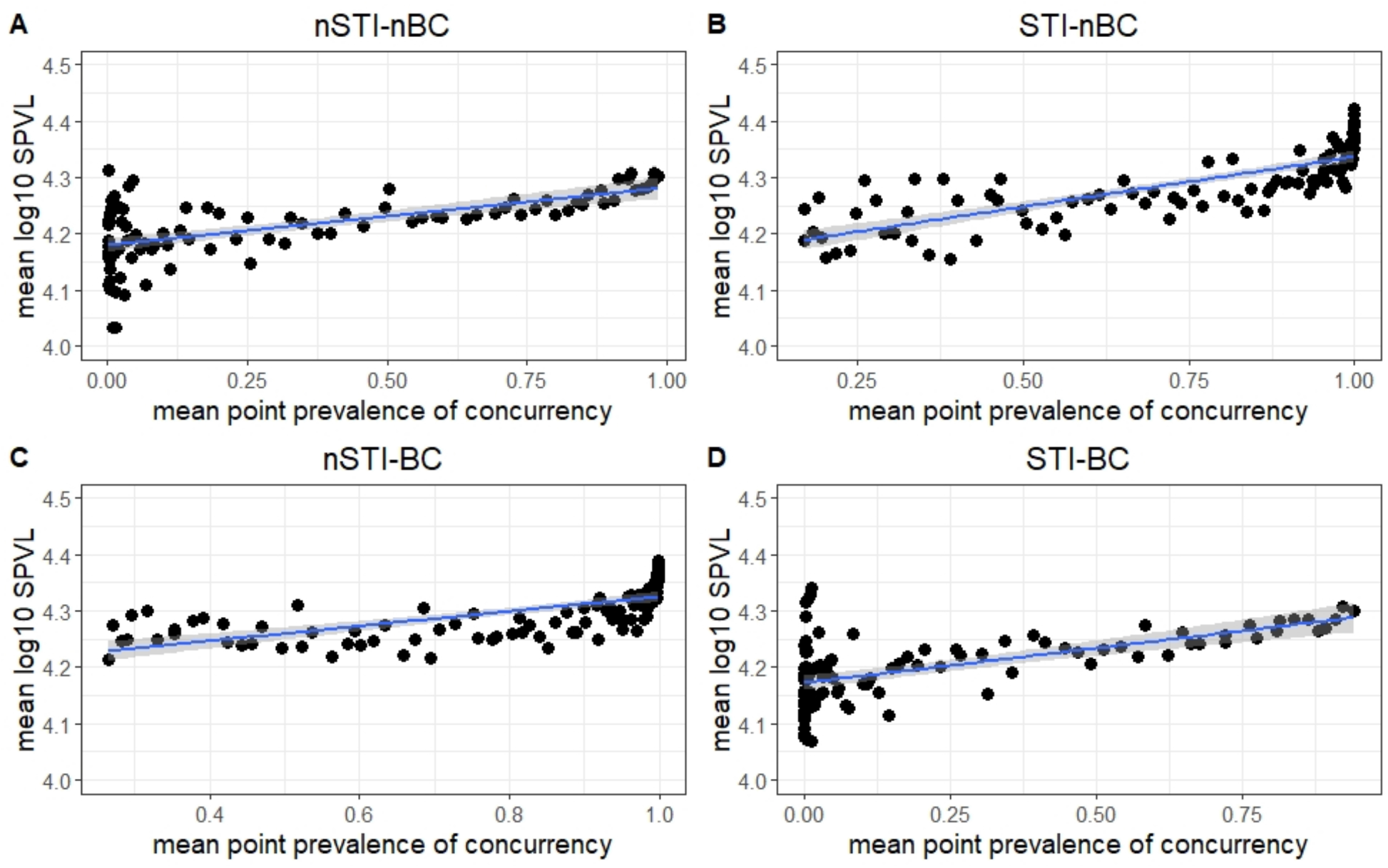
Mean log10 HIV SPVL of all individuals with a date of HIV seroconversion between 1980 and 2015 against mean point prevalence of concurrency over the period 1980-2015 for 100 simulations obtained by varying the weight for the number of partners between −10 and −1 while keeping all other literature and estimated parameters constant at their values in Table S1 and Table 3 respectively (black dots) and the trendline (blue line) with 95% confidence intervals (shaded area). A) no STI co-factor, no behavioural change; B) STI co-factor, no behavioural change; C) no STI co-factor, behavioural change; D) STI co-factor and behavioural change.

## 4 Discussion

After the introduction of ART, increasing trends in HIV SPVL and HIV incidence were observed in MSM [19, 45], which are counter-intuitive and not fully understood. In this study, we used an event- and individual-based model to investigate the impact of STI co-infection, changes in sexual risk behaviour and a combination of both on the evolution of HIV SPVL in MSM.

The results suggest that sexual risk behaviour influences population HIV SPVL (see Figure 4). Furthermore, the model including only an STI co-factor and no behavioural change is the only model that simulates the drop in HIV incidence after an initial increase reported by Van Sighem et al. [45](see Figure 3D). However, the HIV prevalence curves are closer to the observed values in [7] when also including behavioural changes. For the ART coverage, the scenario including only an STI co-factor and no behavioural change results in the closest approximation to the reported value in [20].

In this study, increased access to ART was modeled by increasing the CD4 threshold and assuming earlier diagnosis because of increased HIV testing in the presence of ART. In 2011, the WHO recommended treatment in all serodiscordant couples [51]. However, according to the WHO, many people do not disclose their HIV status with their sexual partner(s) (https://www.who.int/hiv/pub/guidelines/9789241501972/en/). The WHO also reports that to be treated with ART when having CD4 counts higher than the CD4 count initiation threshold, it’s necessary that both partners are tested and are willing to disclose their HIV status to each other. Furthermore the estimated proportion of undiagnosed HIV cases in MSM in 2015 in the Netherlands was about 10% [44]. Therefore, it would not be a realistic scenario to assume that for all serodiscordant couples, the HIV positive partner is on ART. In the model scenarios in this study, about 60% of the serodiscordant couples are on ART. To the best of our knowledge, no estimates are available on the proportion of HIV-positive MSM with CD4 counts higher than the CD4 initiation threshold that are willing to undergo a test and share their HIV status with their partner(s). This hampers making an accurate estimation of the percentage of serodiscordant couples on ART.

Although none of the three scenarios including an STI co-factor and/or behavioural changes could explain the increasing trend in population mean SPVL reported in Gras et al. [19], all scenarios can explain the increase in new HIV cases between 1996 and 2008 reported by Van Sighem et al. [45](see Figure 3D). All scenarios could confirm the higher mean population SPVL with higher relational concurrency reported by Goodreau et al [17](see Figure 4).

There are several potential reasons that none of the scenarios can explain the trends in SPVL reported in Gras et al. [19]. First, in addition to the factors explored in this study (treatment, sexual behaviour and STI co-factor effects), also viral evolution has been reported to have an effect on the population mean SPVL. However, to the best of our knowledge, no modeling framework is available that can incorporate treatment, sexual risk behaviour, STI co-infection and viral evolution into a single model. This will be explored in future research. Second, the available data for this case study as reported in the literature is scarce. Making more data available will lead to better estimation of the model parameters, and possibly to improved models that can better explain the trends in SPVL observed in the literature. Furthermore, the data on SPVL from [19] (see Table 1) for 1995 are based on a considerably lower amount of persons than for 1985 and 2007, which creates uncertainty about the observed trends. According to Cumming and Finch [6], methods for significance testing based on confidence intervals (CI) are reliable when both sample sizes are equal or larger than 10, and when the margins of error differ not more than a factor 2. For 1995 the figure with the data of the mean HIV-1 RNA concentration at set-point in Gras et al. [19] displays less than 10 data points. This would mean that although a proportion overlap of 0.387 between the CIs for 1985 and 1995 is obtained, pointing to a significant difference between the two samples according to Cumming and Finch [6], there is uncertainty about the observed decrease between these two time points. However, the values of the mean log10 SPVL for 1985, 1995 and 2007 in Gras et al. [19] were obtained by fitting a cubic spline through the data of the mean HIV-1 RNA concentration at set-point of 612 MSM with date of seroconversion between 1985 and 2007. Third, only a generic STI co-factor effect was included in the model. Including a more specific STI co-factor effect, e.g. a herpes simplex virus (HSV-2) co-factor effect where all stages of HSV-2 are described in detail may lead to improved models for this case study. However, such model has many more parameters than the model used in this study, and as a consequence requires more data than currently available for parameter estimation.

Weiss et al. [49] implemented a model that fully parametrizes the effect of an STI on HIV transmission to study chlamydia and gonorrhea screening and treatment. However, the model doesn’t implement the effect of HIV on STI transmission, while Simpact Cyan 1.0 does. Furthermore, the model of Weiss et al. is implemented in discrete time, while Simpact Cyan 1.0 is implemented in continuous time. Moreover, Simpact Cyan 1.0 also implements CD4 counts, an option that is not available in the model of Weiss et al.

The results of the model simulations in this study identify factors that should be monitored during future research on the evolution of population mean SPVL. These factors include viral evolution, partnership disclosure and natural history of STI’s. In this way, the models can contribute to the study design of future studies. An overview of previous applications of mathematical models to inform study design in epidemiology is provided by Herzog et al. [25].

To implement a model that fully parametrizes the natural history of an STI, and the effect of this STI on HIV transmission and vice versa, we recommend to collect the following data in future studies:

- monitoring of a cohort of HIV mono-infected, STI mono-infected and HIV-STI co-infected individuals for the study period of interest; collect prevalence and incidence data at least on a yearly basis;
- collecting characteristics for the cohort such as estimated time of HIV infection, estimated time of STI infection and period of HIV ART treatment if applicable.

In summary, the results of this study suggest that sexual risk behaviour could influence SPVL and indicate that more data are needed to test the effect of STI co-factors on SPVL. Furthermore, the trends described in the literature cannot be explained by sexual risk behaviour and STI co-factors only. Future research to understand SPVL evolution should also consider models that include factors related to viral evolution and describe STI co-factors in more detail. To accomplish these goals, more data has to be collected and made available.

## Data Availability

This study concerns a study on mathematical modeling of HIV/AIDS, and uses only information from the literature. It does not include any newly generated data sets or data extracted from repositories.

## Supporting Information

See supplementary.pdf

## Acknowledgments

The research conducted by DMH and NH in this study was funded by the Fonds Wetenschappelijk Onderzoek - Vlaanderen (Research Foundation – Flanders; FWO, http://www.fwo.be/en/) (Grant agreements G0E8416N and G0B2317N).

WD was supported by a postdoctoral fellowship from FWO (12L5816N).

The computational resources and services used in this work were provided by the VSC (Flemish Supercomputer Center), funded by the Research Foundation - Flanders (FWO) and the Flemish Government – department EWI.

We thank Cécile Kremer (I-Biostat,Data Science Institute, UHasselt) for her support on the literature study to obtain the parameter settings described in Table S1 of the Supplementary Material.

