## Supplementary Material for "Influence of sexual risk behaviour and STI co-infection dynamics on the evolution of HIV set point viral load in MSM"

#### Simpact Cyan 1.0

The models in this study were developed with Simpact Cyan 1.0, an open-source framework for individual-based modeling of transmission, diagnosis and treatment of HIV [10]. Simpact Cyan 1.0 models each individual in a population, as well as the sexual relationships between individuals. Birth, mortality, the formation and break-up of relationships as well as HIV transmission, diagnosis and treatment are represented by events. The risk that an event takes place at a certain moment is represented by its hazard function. Simpact Cyan 1.0 implements models in continuous time, i.e. each time an event happens the models are updated. Also a generic STI co-factor effect on HIV is implemented. Furthermore, parameters can be changed during the simulation to simulate interventions. More details on Simpact Cyan 1.0 are available in [11] and from http://www.simpact.org/.

#### Weibull distribution

In Simpact Cyan 1.0, birth events happen as a result of a conception event. As a consequence, no birth events can happen in an MSM population. This means that no new individuals can enter the population after initialization of the simulation. Therefore, the following alternative implementation is used (see Figure S1):

- People are assumed to be born when they have a simulation age of 30 years. In this way, all people with simulation age $< 30$ at initialization enter the population during the simulation.
- People become sexually active at a simulation age of 45 years, corresponding to a real age of 15 years.
- The average simulation age for natural mortality is 110 years, corresponding with a real age of 80 years.

In this way, we avoid that the population decreases during the simulation due to natural (non-AIDS) mortality.

The population pyramid for males from Belgium in 1980 (`https://www.populationpyramid.net/belgium/1980/`) was used to fit the Weibull survival distribution. Figure S2 shows the survival distribution for real age and simulation age.

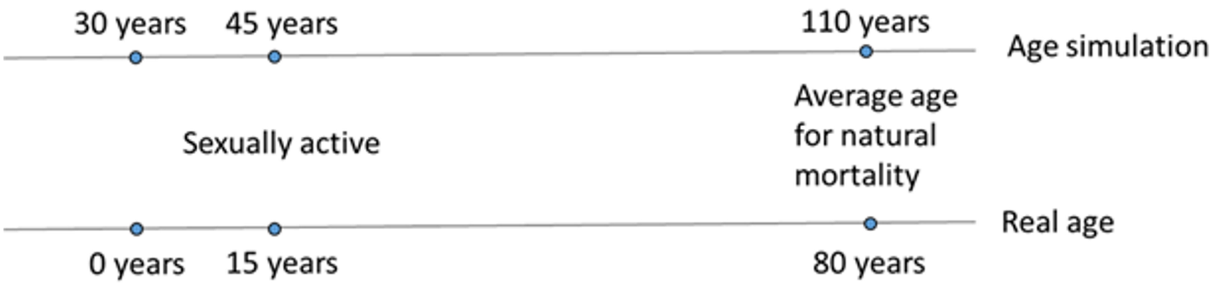


**Figure S1.** Real age versus simulation age in this study. The figure shows birth (30 = simulation age, 0 = real age), the age at with a person becomes sexually active (45 = simulation age, 15 =real age) and the average age for natural mortality, used for fitting the Weibull distribution in Figure S2 (110 = simulation age, 80=real age).

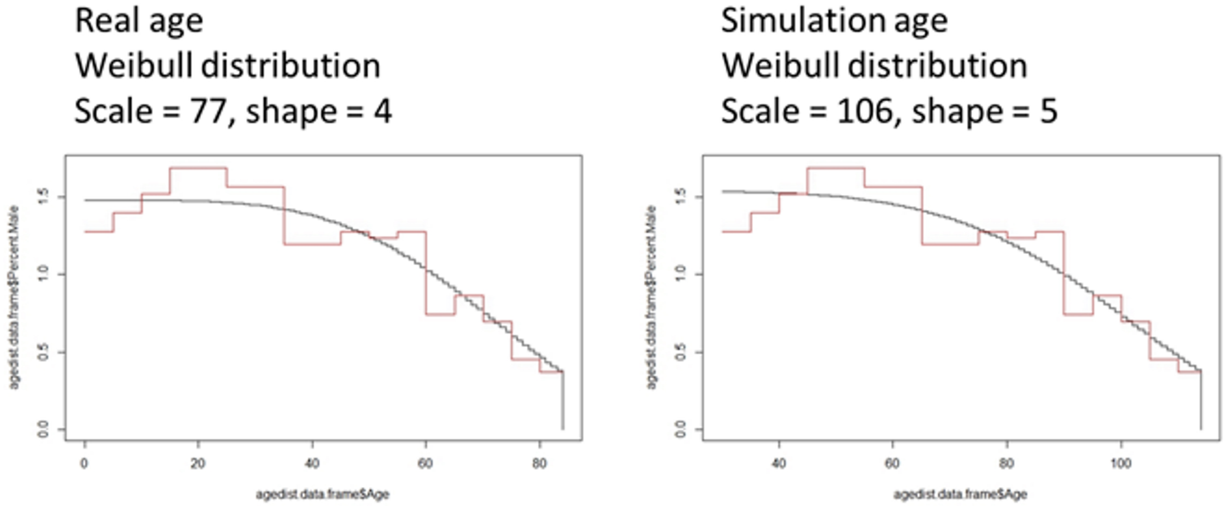


**Figure S2.** Weibull survival distribution for real age (left) and simulation age (right).

In the parameter configuration file, this is implemented as

- mortality.normal.weibull.scale = 106
- mortality.normal.weibull.shape = 5

#### Other parameter settings

**Table S1.** Parameter settings used for all scenarios in this study. The number of parameters is the same in all models. Adding or removing parameters from the model is done by setting parameters to a value so that effects in the model are disabled or not. The parameter names in the first column are directly related to Simpact Cyan 1.0.

| parameter | value | explanation |
|---|---|---|
| mortality.normal.weibull.genderdiff | 0 | no females in the population |
| population.msm | yes | homosexual population |

| | | |
|---|---|---|
| debut.debutage | 45 | 45 = simulation age, 15 = real age (default) |
| population.simtime | 45 | stabilization period of 10 years + 1980 − 2015 |
| population.nummen | 10,000 | population of 10,000 men |
| population.numwomen | 0 | no women in the population |
| population.maxevents | population.simtime × population.nummen × 15 | The average number of events per year that can happen to the same person is 15. The simulation is stopped when a number of events equal to population.maxevents is executed. |
| population.eyecap.fraction | 0.2 | Each men can have sexual contacts with 20% of all other men. Because of age-mixing etc, this will be only max. a few 100 in practice. |
| hsv2transmission.hazard.d | 0.76 in case of an STI-cofactor; not used in case of no STI co-factor | effect of HIV on STI transmission, for HSV2 risk ratio = 2.14 [9]; $d = ln(2.14) = 0.76$ |
| hivseed.time | 10 | stabilization period of 10 years |
| hivseed.type | amount | a fixed number of seeders is chosen to start the HIV infection |
| hivseed.age.min | 45 | 45 = simulation age, 15 = real age (equal to the default debut age) |
| hivseed.age.max | 80 | 80 = simulation age, 50 = real age |
| hivseed.gender | male | HIV seeders are MSM |
| hivseed.amount | 30 | seed of 1% in 1980; start of HIV in MSM end 70's - begin 80's [13]. In 1980, about 3000 of of 10,000 men in the simulation have reached the sexual debut age of 15 (simulation age 45 years). 1% of 3000 = 30 HIV seeds. |
| hsv2seed.time | 10 in case of an STI co-factor; -1 in case of no STI co-factor to disable HSV2 seeding | stabilization period of 10 years |

| | | |
|---|---|---|
| hsv2seed.type | amount in case of an STI co-factor; not used in case of no STI co-factor | a fixed number of seeders is chosen to start the STI infection |
| hsv2seed.age.min | 45 in case of an STI co-factor; not used in case of no STI co-factor | 45 = simulation age, 15 = real age (equal to the default debut age) |
| hsv2seed.age.max | 80 in case of an STI co-factor; not used in case of no STI co-factor | 80 = simulation age, 50 = real age |
| hsv2seed.gender | male in case of an STI co-factor; not used in case of no STI co-factor | STI seeders are MSM |
| hsv2seed.amount | 330 in case of an STI co-factor; not used in case of no STI co-factor | 11% men with HSV2 in 1976 [6]. In 1980, about 3000 of of 10,000 men in the simulation have reached the sexual debut age of 15 (simulation age 45 years). 11% of 3000 = 330 HSV2 seeds. |
| hivtransmission.param.e1 | 0.4700036 | risk ratio = 1.6; ln(1.6)=0.4700036 [16] |
| hivtransmission.param.e2 | 0.4700036 | risk ratio = 1.6; ln(1.6)=0.4700036 [16] |
| person.hsv2.a.dist.type | fixed in case of an STI co-factor; not used in case of no STI co-factor | person dependent baseline value for the STI transmission hazard is a fixed value |
| person.hsv2.a.dist.fixed.value | -2.25 in case of an STI co-factor; not used in case of no STI co-factor | transmission rate (cumulative incidence) = 10% [4]; risk ratio = -log(1 − cumulative incidence) = -log (0.9) = 0.1053605; ln(0.1053605) = -2.25 |
| mortality.aids.survtime.C | 65 | parameter C in the formula for survival time [2] $t_{survival} = (C/V_{sp}^k) \times 10^x$ where $V_{sp}$ is the set-point viral load and x is the parameter determined per person allowing some randomness in the formula; results in survival time of 10-11 years for AIDS [18] |

| | | |
|---|---|---|
| mortality.aids.survtime.k | -0.2 | parameter k in the formula for survival time [2] (see parameter C); results in survival time of 10-11 years for AIDS [18] |
| person.survtime.logoffset.dist.type | normal | the parameter x in the formula for survival time is drawn from a normal distribution |
| person.survtime.logoffset.dist.normal.mu | 0 | the normal distribution for parameter x in the formula for survival time has mean 0 |
| person.survtime.logoffset.dist.normal.sigma | 0.1 | the normal distribution for parameter x in the formula for survival time has standard deviation 0.1; to obtain a survival distribution which is approximately Weibull distributed |
| person.vsp.model.logdist2d.dist2d.binormalsymm.mean | 4.3 | the distribution used to pick set-point viral load values on a base 10 logarithmic scale is a binormal distribution using the same mean and standard deviation for the x-direction as for the y-direction (default); the parameters are fitted to the histogram for MSM in [7]; the mean is 4.3 |
| person.vsp.model.logdist2d.dist2d.binormalsymm.rho | 0.31 | parameter rho for the correlation between x and y in the distribution of set-point viral load; this parameter represents set-point viral load heritability and is taken from [3] |
| person.vsp.model.logdist2d.dist2d.binormalsymm.sigma | 1.3 | standard deviation for the distribution of set-point viral load; the parameters are fitted to the histogram for MSM in [7] |
| person.vsp.model.logdist2d.usealternativeseeddist | yes | to use another distribution than the default to initialize set-point viral load values when HIV seeding is triggered |

| | | |
|---|---|---|
| person.vsp.model.logdist2d.alternativeseed.dist.type | fixed | person dependent initial set-point viral load value is a fixed value |
| person.vsp.model.logdist2d.alternativeseed.dist.fixed.value | 4 | value is a bit lower than the value for 1984 in [8] |
| formationmsm.hazard.type | simple | amount of data available is limited; we take the most simple form of the formation hazard to limit the amount of parameters to fit |
| person.art.accept.threshold.dist.fixed.value | 1 | ART acceptance threshold is 1 (the maximum value for this parameter) for each person; willingness to accept treatment when offered. RIVM reports that in 2017, 95% of MSM diagnosed and linked to care were on ART [15] |
| diagnosis.baseline | -1000000 | no ART before 1994 [12]; we set this parameter to a very small number so that nobody will be treated |
| person.cd4.start.dist.type | lognormal | lognormal distribution for the CD4 value a person has at the time of infection |
| person.cd4.start.dist.lognormal.zeta | $ln\left(\frac{mu.cd4}{\sqrt{\frac{1+var.cd4}{mu.cd4^2}}}\right)$ | value for location parameter zeta of the distribution for the CD4 value a person has at the time of infection; mu.cd4=800; var.cd4=40000 |
| person.cd4.start.dist.lognormal.sigma | $\sqrt{ln\left(\frac{1+var.cd4}{mu.cd4^2}\right)}$ | value for scale parameter sigma of the distribution for the CD4 value a person has at the time of infection; mu.cd4=800; var.cd4=40000 |
| person.cd4.end.dist.type | lognormal | lognormal distribution for the CD4 value a person will have when he dies from AIDS related causes |
| person.cd4.end.dist.lognormal.zeta | $log\left(\frac{mu.cd4.end}{\sqrt{\frac{1+var.cd4.end}{mu.cd4.end^2}}}\right)$ | value for location parameter zeta of the distribution for the CD4 value a person will have when he dies from AIDS related causes; mu.cd4.end=20; var.cd4.end=5 |

| | | |
|---|---|---|
| person.cd4.end.dist.lognormal.sigma | $\sqrt{log\left(\frac{1+var.cd4.end}{mu.cd4.end^2}\right)}$ | value for scale parameter sigma of the distribution for the CD4 value a person will have when he dies from AIDS related causes; mu.cd4.end=20; var.cd4.end=5 |

### Intervention events

Table S2 summarizes the intervention events for simulating the increased ART coverage since 1994, and the increase in sexual risk behaviour with ART coverage.

**Table S2.** Intervention events for simulating the increased ART coverage since 1994, and the increase in sexual risk behaviour with ART coverage. CD4 treshold: the parameter monitoring.cd4.threshold, if a person's CD4 count is below this threshold, he will be offered ART; diagnosis.baseline: baseline value in the hazard for a diagnosis event; formation alpha_12: the parameter formationmsm.hazard.simple.alpha_12 in the hazard for a relationship formation event, corresponding to a weight for the number of relationships men in the relationship have. This parameter only changes for the scenarios with increased risk behaviour. The behavioural parameter formation alpha_12 is equal to an initial value in the period 1980-1993. From 1994 onwards until 2014, this value is increased with 0.05 every two years to simulate increased risk behaviour over time in the decade after the introduction of ART. The actual value of the initial value of formation alpha_12 is determined during the model calibration procedure.

| **year** | **time** | **CD4 threshold** | **diagnosis.baseline** | **formation alpha_12** |
|---|---|---|---|---|
| 1994 | 24 | 200 | -1.5 | initial value + 0.05 |
| 1996 | 26 | 230 | -1.25 | initial value + 0.1 |
| 1998 | 28 | 260 | -1 | initial value + 0.15 |
| 2000 | 30 | 290 | -0.75 | initial value + 0.2 |
| 2002 | 32 | 320 | -0.5 | initial value + 0.25 |
| 2004 | 34 | 350 | -0.25 | initial value + 0.3 |
| 2006 | 36 | 380 | 0 | initial value + 0.35 |
| 2008 | 38 | 410 | 0.25 | initial value + 0.4 |
| 2010 | 40 | 440 | 0.5 | initial value + 0.45 |
| 2012 | 42 | 470 | 0.75 | initial value + 0.5 |
| 2014 | 44 | 500 | 1 | initial value + 0.55 |

### Statistical analysis of the parameter space

#### Methods

For exploring the parameter space, we consider the 10,000 parameter combinations generated by Latin Hypercube Sampling (LHS) and focus on the top 1% of the lowest values of the GOF statistic (sum of squared relative errors) and follow the approach described by Castro Sanchez et al. [5]. First, for each parameter, smoothed density plots of the initial uniform distribution (range as in Table 2) are compared with smoothed density plots for the distribution of the top 1% solutions. The more peaked the density for the top 1% solutions, the more the parameter is influenced by the data. Second, a classification tree method, called activity region finder (ARF) [1] is applied to identify regions in the parameter space that have significantly more top 1% solutions than other solutions. The 10,000 parameter combinations from the LHS are used as input variables, and a binary output, which is 1 for a top 1% solution and 0 otherwise is used. Third, the input-output from the ARF is used to fit generalized additive models (GAM) [17]. For each parameter, the predicted probability of low sum of relative squared errors is plotted against the value of the parameter. In this way, we further explore which regions in the parameter space contain a large amount of top 1% solutions. Finally, the associations between the parameters in the top 1% solution space are calculated using the Maximal Information Coefficient (MIC) [14].

#### Model with no STI co-factor and no behavioural change (nSTI-nBC)

The distribution of the top 1% solutions showed the most pronounced peaks for the weight for the age of the parameters (formation $\alpha_4$) and the parameter c in the HIV transmission hazard (HIV transmission c), followed by the weight for the number of partners (formation $\alpha_{12}$) and its assortativity (formation $\alpha_3$)(see Figure S3).

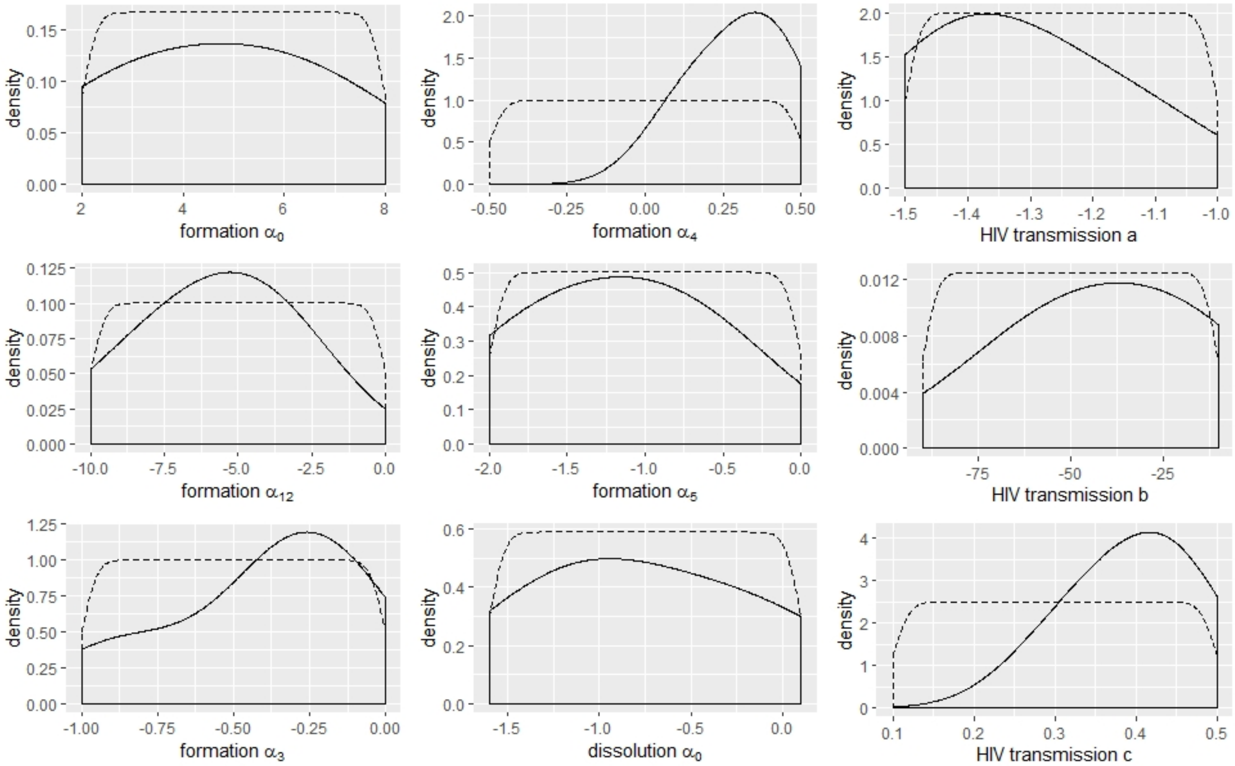


**Figure S3.** Analysis of the parameter space for the nSTI-nBC model. Smoothed density plots for the initial uniform distribution (dashed line) and the distribution of the top 1% solutions (solid line) for the nine estimated parameters.

When applying ARF, only the weight for the age of the partners (formation $\alpha_4$), the assortativity for the number of partners (formation $\alpha_3$), the baseline dissolution parmeter (dissolution $\alpha_0$) and the assortativity for age (formation $\alpha_5$) take part in the classification process. The first split of the classification tree uses the weight for the age of the partners (formation $\alpha_4$). The next splits use the assortativity for the number of partners (formation $\alpha_3$) and the baseline dissolution parameter (dissolution $\alpha_0$). Only one region with significantly more top 1% solutions could be detected, where the weight for the age of the partners (formation $\alpha_4$) is between 0.3917 and 0.4485.

The results of the GAM (Figure S4) show a high probability of low relative sum of squared errors (SSE) for positive values of the weight for the age of the partners (formation $\alpha_4$). Lower values of the baseline formation parameter (formation $\alpha_0$), the weight for the number of partners (formation $\alpha_{12}$), the assortativity for age (formation $\alpha_5$) and the baseline parameter for HIV transmission (HIV transmission a) increase the probability of low relative SSE. Higher values of the HIV transmission parameters related to viral load (HIV transmission b and c) increase the probability for low relative SSE.

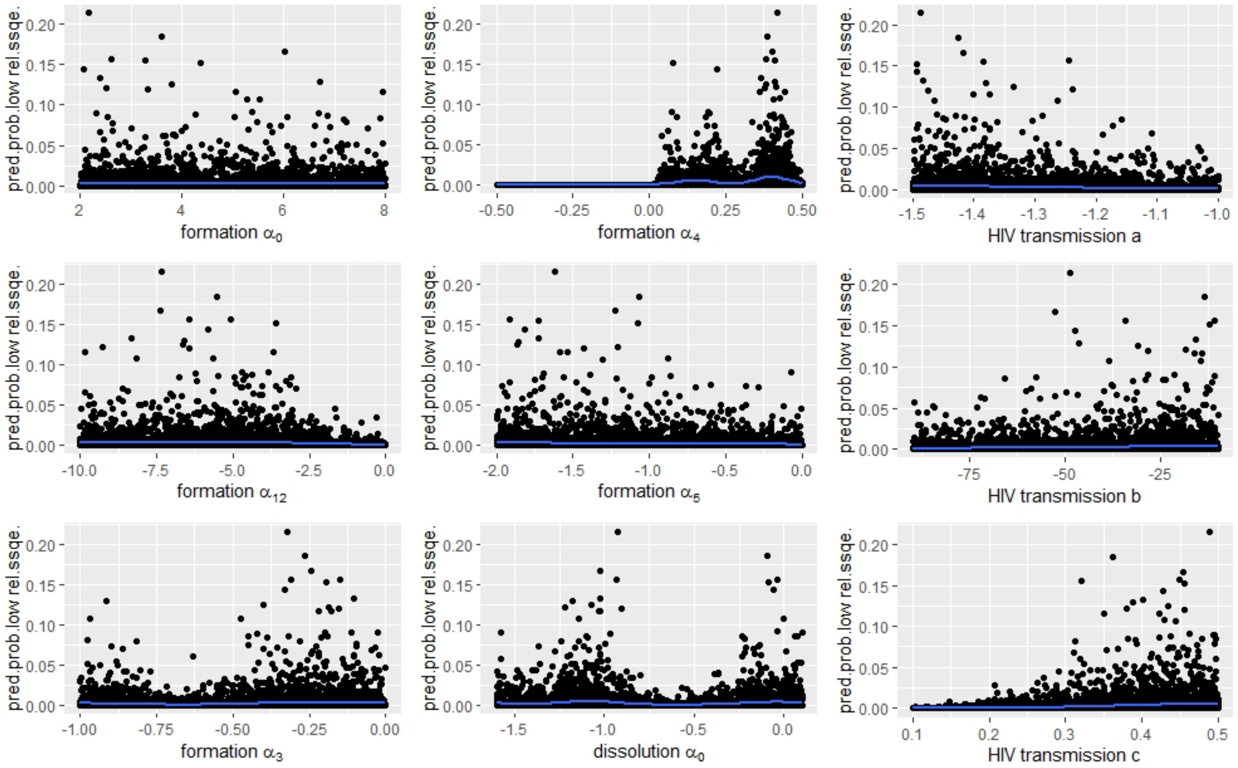


**Figure S4.** Analysis of the parameter space for the nSTI-nBC model. Results of the GAM: predicted probabilities of low relative sum of squared errors (SSE) for the nine estimated parameters.

The three highest values for the MIC were obtained for

1. the weight for the number of partners (formation $\alpha_{12}$) and the weight for the age of the partners (formation $\alpha_4$)(MIC = 0.71, negative correlation);

2. the weight for the number of partners (formation $\alpha_{12}$) and the baseline dissolution parameter (dissolution $\alpha_0$)(MIC = 0.53, positive correlation);

3. the weight for assortativity for the number of partners (formation $\alpha_3$) and the baseline dissolution parameter (dissolution $\alpha_0$)(MIC = 0.52, positive correlation).

#### Model with an STI co-factor and no behavioural change (STI-nBC)

The distribution of the top 1% solutions showed the most pronounced peaks for the weight for the age of the partners (formation $\alpha_4$) and the parameter c in the HIV transmission hazard (HIV transmission c), followed by the baseline parameter a in the HIV transmission hazard (HIV transmission a)(see Figure S5).

When applying ARF, only the parameter c in the HIV transmission hazard (HIV transmission c), the weight for the number of partners (formation $\alpha_{12}$), the weight for the age of the partners (formation $\alpha_4$) and the weight for the assortativity related to the number of partners (formation $\alpha_3$) take part in the classification process. The first split of the classification tree uses the parameter c in the HIV transmission hazard (HIV transmission c). The next splits use the weight for the number of partners (formation $\alpha_{12}$) and the weight for the age of the partners (formation $\alpha_4$). Only one region with significantly more top 1% solutions could be detected, where the HIV transmission parameter c (HIV transmission c) is between 0.3447 and 0.3481.

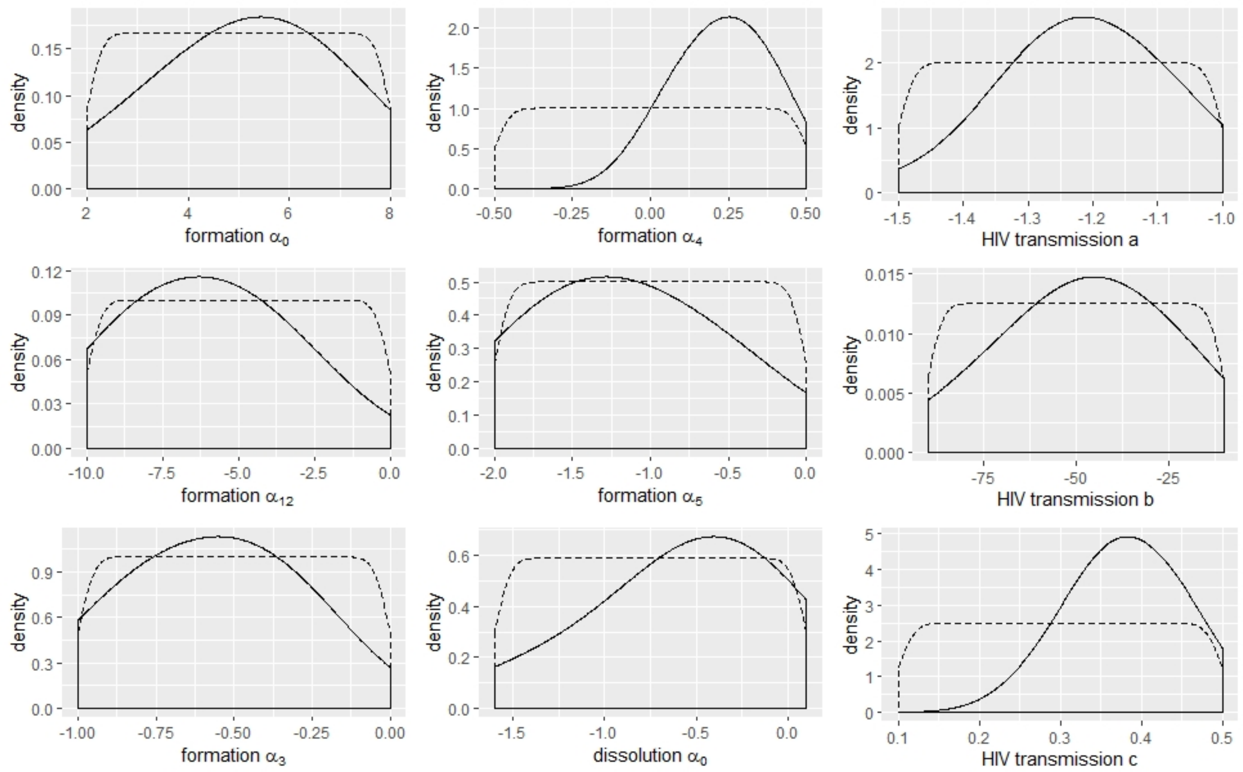


**Figure S5.** Analysis of the parameter space for the STI-nBC model. Smoothed density plots for the initial uniform distribution (dashed line) and the distribution of the top 1% solutions (solid line) for the nine estimated parameters.

The GAM predicted a low relative SSE for positive values of the weight for the age of the partners (formation $\alpha_4$) and values between 0.2 and 0.5 for the HIV transmission parameter c (HIV transmission c)(see Figure S6). Lower values of the weight for the number of partners (formation $\alpha_{12}$), the assortativity related to the number of partners (formation $\alpha_3$) and the assortativity related to the age of the partners (formation $\alpha_5$) increase the probability for a low relative SSE. Higher values of the baseline dissolution parameters (dissolution $\alpha_0$ increase the probability for a low relative SSE.

The three highest MIC values were obtained for

1. the weight for the number of partners (formation $\alpha_{12}$) and the weight for the age of the partners (formation $\alpha_4$)(MIC = 0.58, negative correlation);

2. the weight for assortativity related to the number of partners (formation $\alpha_3$) and the weight for assortativity related to the age of the partners (formation $\alpha_5$)(MIC = 0.45, positive correlation);

3. the weight for the age of the partners (formation $\alpha_{12}$) and the HIV transmission parameter c (HIV transmission c)(MIC = 0.42, negative correlation).

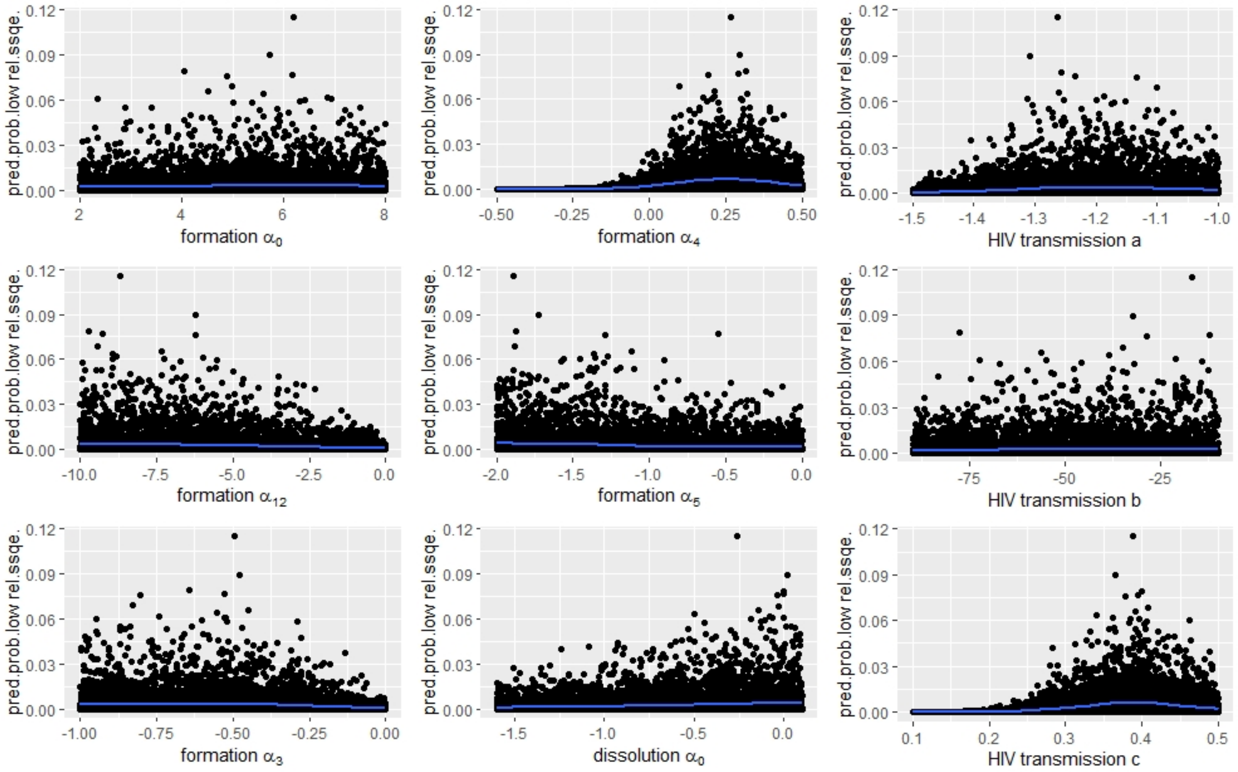


**Figure S6.** Analysis of the parameter space for the STI-nBC model. Results of the GAM: predicted probabilities of low relative sum of squared errors (SSE) for the nine estimated parameters.

#### Model with no STI co-factor and a behavioural change (nSTI-BC)

The distribution of the top 1% solutions showed the most pronounced peak for the weight for the age of the partners (formation $\alpha_4$), followed by the HIV transmission parameters b and c related to viral load (HIV transmission b and c)(see Figure S7).

When applying ARF, only the weight for the age of the partners (formation $\alpha_4$) and the weight for the number of partners (formation $\alpha_{12}$) take part in the classification process. The first split of the tree uses the weight for the age of the partners (formation $\alpha_4$). The next splits use the weight for the number of partners (formation $\alpha_{12}$). No regions with significantly more top 1% solutions could be detected.

The results of the GAM (Figure S8) show that positive values of the weight for the age of the partners (formation $\alpha_4$) are associated with a low relative SSE. Higher values of the HIV transmission parameters related to viral load (HIV transmission b and c) increase the probability for a low relative SSE.

The three highest MIC values were obtained for

1. the weight for the number of partners (formation $\alpha_{12}$) and HIV transmission parameter c (negative correlation; MIC = 0.72);

2. HIV transmission parameters a and c (negative correlation; MIC = 0.62);

3. the weight for the number of partners (formation $\alpha_{12}$) and the weight for the age of the partners (negative correlation; MIC = 0.61).

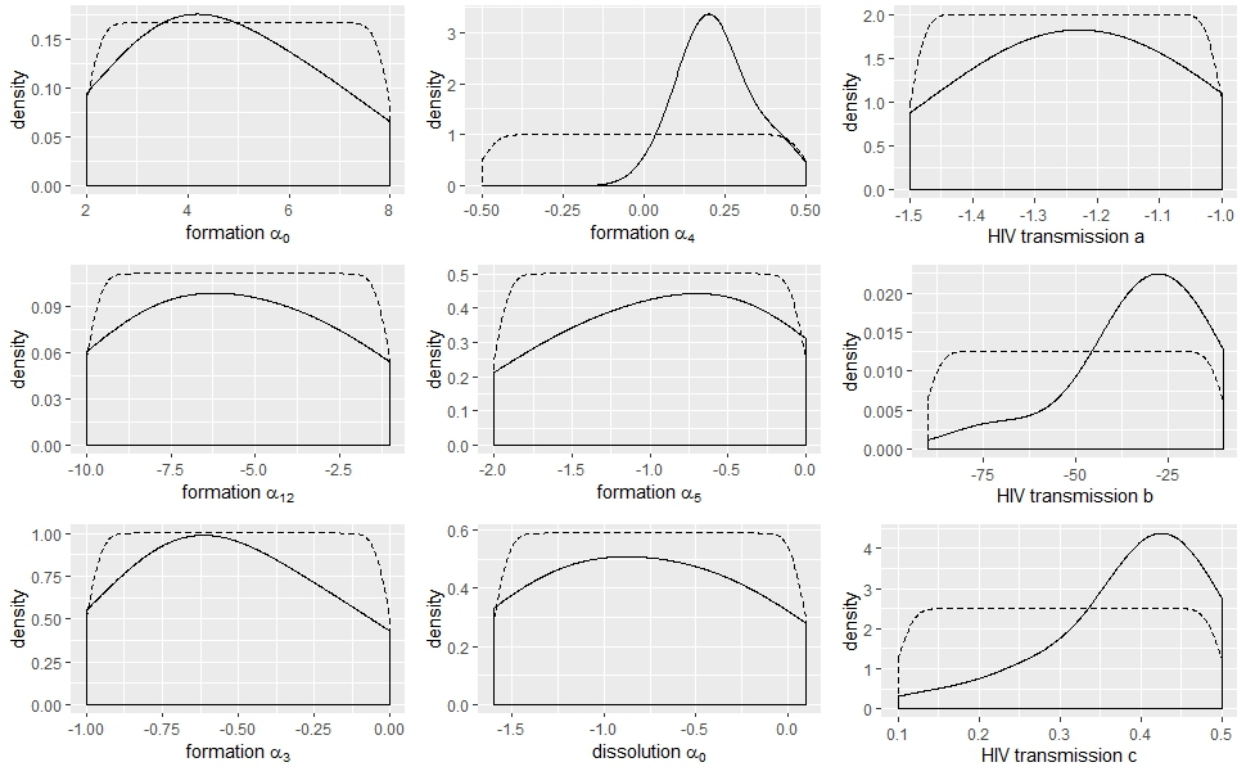


**Figure S7.** Analysis of the parameter space for the nSTI-BC model. Smoothed density plots for the initial uniform distribution (dashed line) and the distribution of the top 1% solutions (solid line) for the nine estimated parameters.

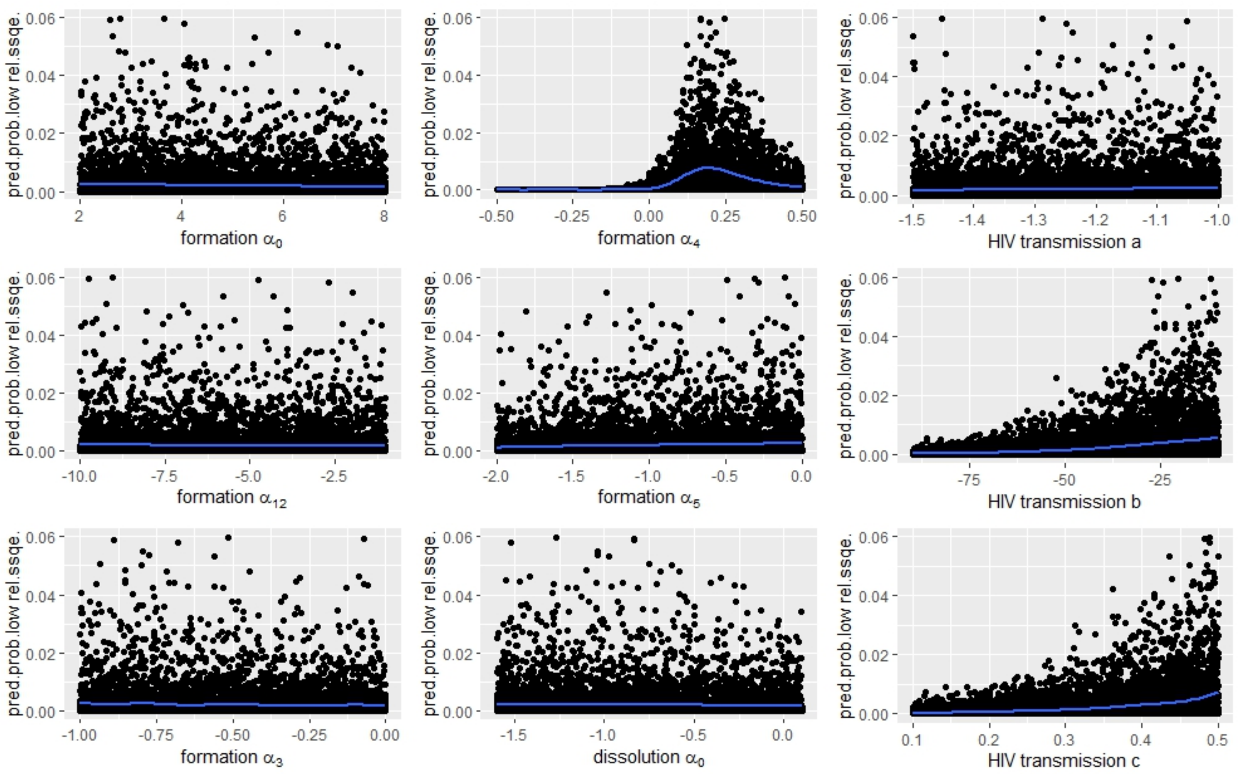


**Figure S8.** Analysis of the parameter space for the nSTI-BC model. Results of the GAM: predicted probabilities of low relative sum of squared errors (SSE) for the nine estimated parameters.

#### Model with an STI co-factor and a behavioural change (STI-BC)

The distribution of the top 1% solutions showed the most pronounced peaks for the weight for the age of the partners (formation $\alpha_4$) and the HIV transmission parameter c (HIV transmission c), followed by the baseline parameter for relationship formation (formation $\alpha_0$)(see Figure S9.

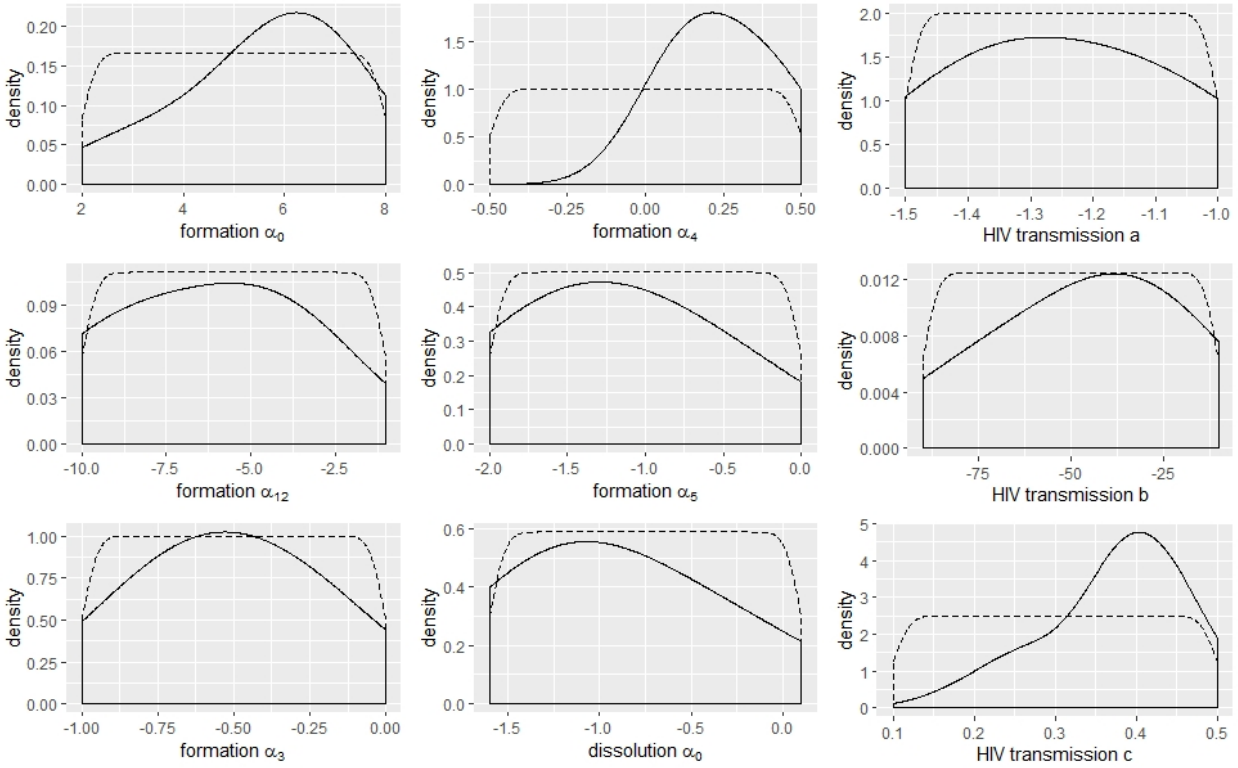


**Figure S9.** Analysis of the parameter space for the STI-BC model. Smoothed density plots for the initial uniform distribution (dashed line) and the distribution of the top 1% solutions (solid line) for the nine estimated parameters.

When applying ARF, only the weight for the age of the partners (formation $\alpha_4$), the weight for the number of partners (formation $\alpha_{12}$) and HIV transmission parameter a take part in the classification process. The first split of the tree uses the weight for the age of the partners (formation $\alpha_4$). The next splits use the weight for the number of partners (formation $\alpha_{12}$) and HIV transmission parameter a. Two regions with significantly more top 1% solutions could be detected. In the first region the weight for the age of the partners (formation $\alpha_4$) is between 0.22686 and 0.23041. In the second region the weight for the age of the partners (formation $\alpha_4$) is between -0.4999 and 0.2268 and the weight for the number of partners (formation $\alpha_{12}$) is between -3.497 and -3.465.

The results of the GAM (Figure S10) show that positive values of the weight for the age of the partners (formation $\alpha_4$) are associated with a low relative SSE. Furthermore, higher values of the baseline formation parameter (formation $\alpha_0$) and the HIV transmission parameters related to viral load (HIV transmission b and c) increase the probability for a low relative SSE. Lower values for the weight for assortativity related to the age of the partners (formation $\alpha_5$) and the baseline dissolution parameter (dissolution $\alpha_0$) increase the probability for a low relative SSE. For the weight for the number of partners (formation $\alpha_{12}$), two regions with higher probability of low relative SSE were observed: a first region where $\alpha_{12}$ has values between -10 and -8.75, and a second region where $\alpha_{12}$ has values between -6.25 and -2.5.

The three highest MIC values were obtained for

1. the HIV transmission parameters b and c related to viral load (MIC = 0.65, negative correlation);

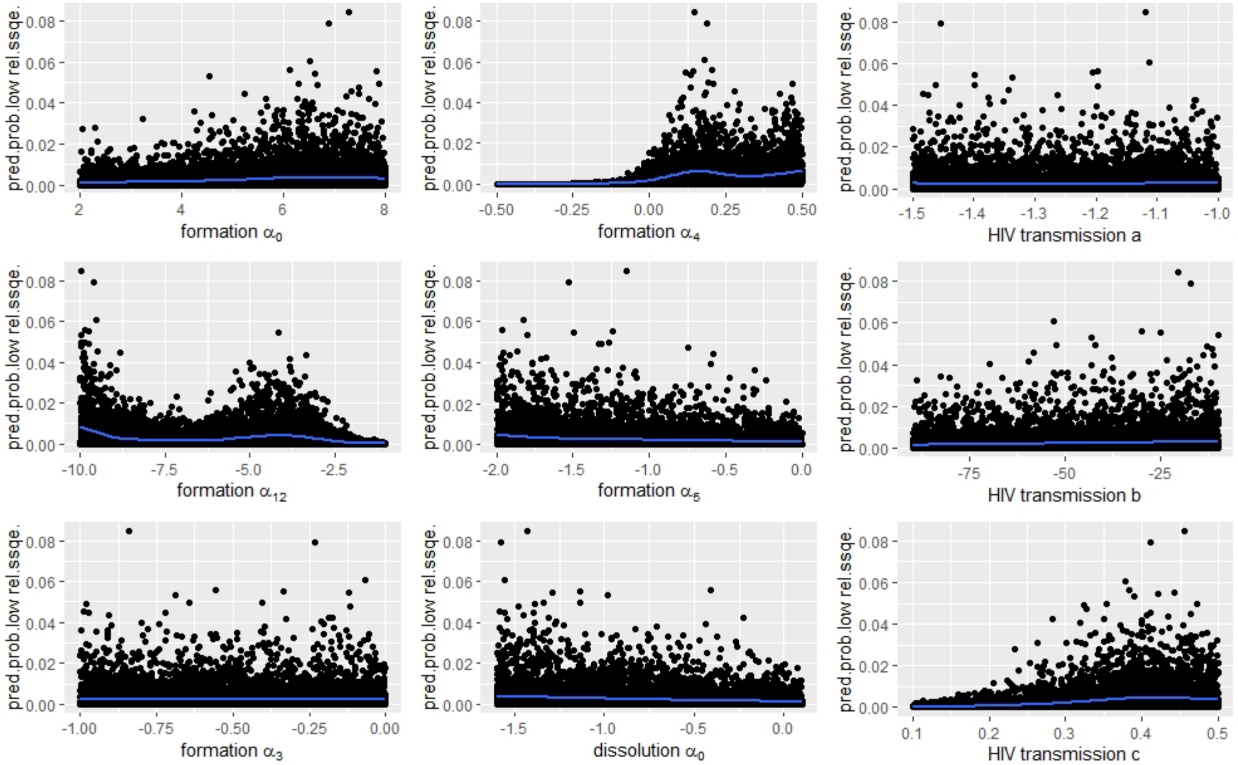


**Figure S10.** Analysis of the parameter space for the STI-BC model. Results of the GAM: predicted probabilities of low relative sum of squared errors (SSE) for the nine estimated parameters.

2. the weight for the number of partners (formation $\alpha_{12}$) and the baseline dissolution parameters (dissolution $\alpha_0$) (MIC = 0.58, negative correlation);

3. the baseline dissolution parameter (dissolution $\alpha_0$) and the HIV transmission parameter c (MIC = 0.57, positive correlation).

#### Generation of Figure 2

For each of the four scenarios (nSTI-nBC, STI-nBC, nSTI-BC, STI-BC) 100 simulations with the fitted parameters were run. For each simulation, and for each person with date of seroconvertion between 1980-2015, we determined the first HIV RNA at 9-27 months after seroconvertion (red dots in Figure S11, each dot represents a person). A curve of the mean log10 SPVL (black line in Figure S11 over time was generated by fitting a cubic spline through the red dots, in analogy to Figure 1A in Gras et al [8]. This was repeated for all 100 simulations. Figure 2 in the main text represents the median and interquartile range of the 100 splines.

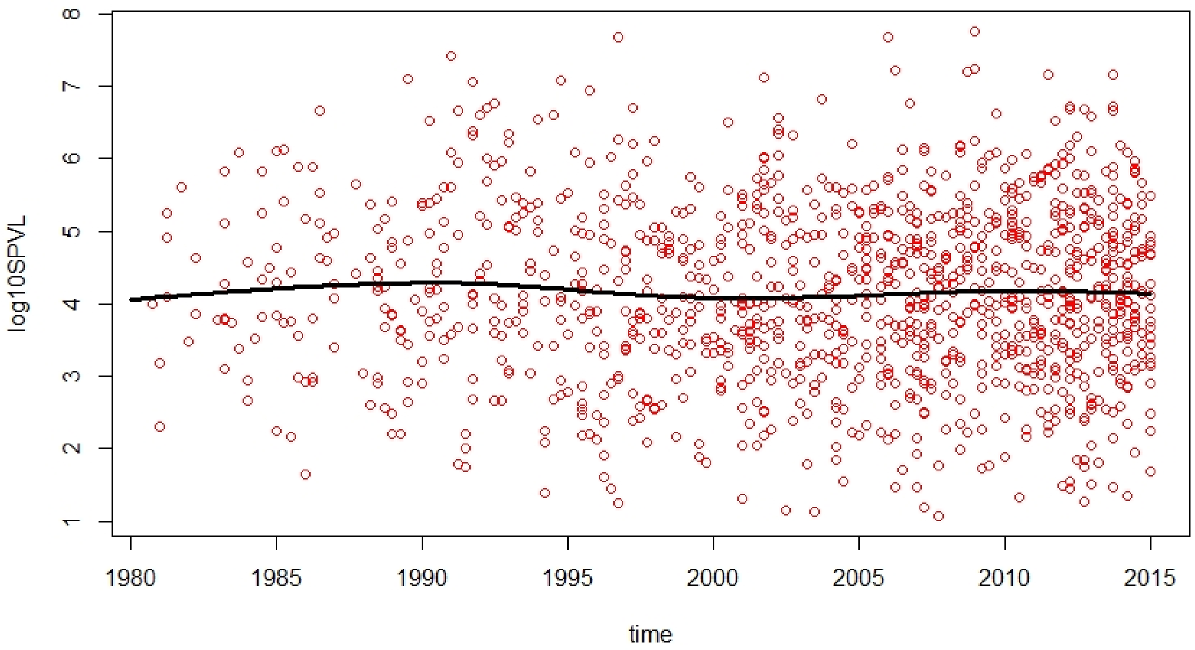


**Figure S11.** First HIV RNA at 9-27 months after seroconversion for each person with date of seroconversion between 1980 and 2015 (red dots) for a single simulation. Black line: cubic spline fitted through the red dots, representing the mean log10 SPVL over time.

18. World Health Organization. *AIDS epidemic update: December 2007*. WHO Regional Office Europe, 2008.
